# Improving breast cancer diagnostics with artificial intelligence for MRI

**DOI:** 10.1101/2022.02.07.22270518

**Authors:** Jan Witowski, Laura Heacock, Beatriu Reig, Stella K. Kang, Alana Lewin, Kristine Pyrasenko, Shalin Patel, Naziya Samreen, Wojciech Rudnicki, Elżbieta Łuczyńska, Tadeusz Popiela, Linda Moy, Krzysztof J. Geras

**Affiliations:** Department of Radiology, New York University Grossman School of Medicine; Center for Advanced Imaging Innovation and Research, New York University; Department of Population Health, New York University Grossman School of Medicine; Center for Data Science, New York University; Department of Computer Science, Courant Institute of Mathematical Sciences, New York University; Vilcek Institute of Graduate Biomedical Sciences, NYU Grossman School of Medicine; Perlmutter Cancer Center, NYU Langone Health; Electroradiology Department, Jagiellonian University Medical College, Kraków, Poland; Chair of Radiology, Jagiellonian University Medical College, Kraków, Poland

## Abstract

Dynamic contrast-enhanced magnetic resonance imaging (DCE-MRI) has a very high sensitivity in detecting breast cancer, but it often leads to unnecessary biopsies and patient workup. In this paper, we used an artificial intelligence (AI) system to improve the overall accuracy of breast cancer diagnosis and personalize management of patients undergoing DCE-MRI. On the internal test set (N=3,936 exams), our system achieved an area under the receiver operating characteristic curve (AUROC) of 0.92 (95% CI: 0.92-0.93). In a retrospective reader study, there was no statistically significant difference between 5 board-certified breast radiologists and the AI system (mean ΔAUROC +0.04 in favor of the AI system). Radiologists’ performance improved when their predictions were averaged with AI’s predictions (mean ΔAUPRC [area under the precision-recall curve] +0.07). Those hybrid predictions also increase interreader agreement (Fleiss’ kappa Δ +0.21 (0.16-0.26)). We demonstrated the generalizability of the AI system using multiple data sets from Poland and the US. In subgroup analysis, we observed consistent results across different cancer subtypes and patient demographics. Using the decision curve analysis, we showed that the AI system can reduce unnecessary biopsies in the range of clinically relevant risk thresholds. This would lead to avoiding benign biopsies in up to 20% of all BI-RADS category 4 patients. Finally, we performed an error analysis, investigating situations where AI predictions were mostly incorrect. This exploratory work creates a foundation for deployment and prospective analysis of AI-based models for breast MRI.

**One Sentence Summary:** We developed and evaluated an AI system for predicting breast cancer in MRI that matches the performance of board-certified radiologists and has the potential to prevent unnecessary biopsies.

## INTRODUCTION

Breast MRI is a highly accurate modality in detecting breast cancer with reported sensitivity of over 80% (1, 2). Traditionally, its use in screening was limited to high-risk patients. Newer evidence supports the role of screening MRI in intermediate-risk and average-risk women (1, 3, 4). Diagnostic MRI is also useful for additional indications such as problem-solving and patients with newly diagnosed breast cancer (5, 6). As the number of patients undergoing breast MRI continues to increase, it is important to maintain high specificity and positive predictive value to minimize unnecessary biopsies and follow-up recommendations. In the studies of screening MRI for intermediate- and average-risk women, the positive predictive value of a biopsy recommendation ranged from 19.6 to 35.7 percent (7). This translates to 2-4 benign biopsies performed for every malignant biopsy. There is a need to develop well-tested tools to improve the performance of MRI and increase quality of care. Furthermore, current tools and diagnostic algorithms are rarely personalized. Therefore, there is a need to develop tools that take into consideration clinicians’ or patients’ preferences, for example when deciding on whether to biopsy or not.

Artificial intelligence has had an enormous impact on medical research, including breast imaging. It has been shown to improve breast cancer detection rates in full-field mammography, digital breast tomosynthesis, breast ultrasound, and breast MRI (8–16). While published studies on AI for breast imaging usually demonstrate good classification performance, few measure potential clinical impact. Moreover, evaluation of external and heterogeneous data sets is critical to validate how AI systems may perform in different populations. Providing extensive, patient-oriented results is necessary to gain the trust of practitioners. Furthermore, it builds evidence that enables the translation of results into prospective studies and ultimately, clinical practice.

In our paper, we describe an AI system used to predict the probability of breast cancer in patients undergoing DCE-MRI. We extensively evaluate its performance on internal and international external data sets using both traditional metrics for diagnostic accuracy (area under the receiver operating characteristic curve [AUROC], area under the precision-recall curve [AUPRC], sensitivity, specificity, positive predictive value [PPV] and negative predictive value [NPV]), as well as methods evaluating clinical utility impact on decision making, such as decision curve analysis. We compare AI system’s performance to radiologists in a retrospective reader study. We explore how the system performs in various patient subgroups and examine cases where the AI system failed to produce correct diagnoses. Finally, we analyze a specific clinical scenario — personalizing the management of BI-RADS 4 patients — leading to reduction in unnecessary biopsies.

## RESULTS

### Patient data

Our internal data set used for model training and evaluation includes 21,537 bilateral DCE-MRI studies (N=13,463 patients) who underwent a breast MRI between 2008 and 2020 at one of the NYU Langone Health breast imaging sites. All studies have been performed with either 1.5T magnet or 3T magnet MRI scanners (Table S3). Data included patients reporting for high-risk screening, preoperative planning, routine surveillance, follow-up after suspicious findings in previous studies, and workup of equivocal findings reported in mammography or ultrasound. We excluded patients after bilateral mastectomy, patients after neoadjuvant chemotherapy, and patients with MRI performed to assess implant integrity. T1-weighted fat-saturated pre-contrast and at least two post-contrast series were required for the study to be included in the data set. The entire data set was initially split into training, validation and test subsets with a 60-15-25 ratio. Additional filtering and manual evaluation was performed to ensure consistency of the data set and ground truth labels (see Methods). In addition to DCE-MRI studies, we collected associated radiology and pathology reports, and patients demographic data (Tables 1, 2).

**Table 1:** NYU Langone data set breakdown including demographic data and imaging characteristics. Values are *N* (%) unless specified otherwise. Background parenchymal enhancement (BPE) and the amount of fibroglandular tissue (FGT) are reported according to the American College of Radiology BI-RADS Atlas 5th edition (17). Studies which had missing BI-RADS, BPE and FGT categories are omitted in this table. †Because malignant and benign labels are not mutually exclusive, the total number of malignant, benign and negative studies can be greater than 100%. Breast-level label statistics are presented in Table S4.

|  | Training set | Validation set | Test set | Total |
| --- | --- | --- | --- | --- |
| **Studies** | 14,198 | 3,403 | 3,936 | 21,537 |
| **Patients** | 8,679 | 2,142 | 2,642 | 13,463 |
| **Age**, mean years (SD) | 54.56±13.2 | 54.30±13.0 | 55.23±12.7 | 54.64±13.1 |
| **Race** |  |  |  |  |
| White | 9,819 (69.2) | 2,317 (68.1) | 2,738 (69.6) | 14,874 (69.1) |
| Black | 802 (5.6) | 205 (6.0) | 244 (6.2) | 1,251 (5.8) |
| Asian | 548 (3.9) | 182 (5.3) | 163 (4.1) | 893 (4.1) |
| Other/Unknown | 3,005 (21.2) | 696 (20.5) | 781 (19.8) | 4,482 (20.8) |
| **Label<sup>†</sup>** |  |  |  |  |
| Malignant | 2,337 (16.5) | 582 (17.1) | 861 (21.9) | 3,780 (17.6) |
| Benign | 3,380 (23.8) | 804 (23.6) | 1,148 (29.2) | 5,332 (24.8) |
| Negative | 10,040 (70.7) | 2,397 (70.4) | 2,491 (63.3) | 14,928 (69.3) |
| **BI-RADS category** |  |  |  |  |
| BI-RADS 0 | 446 (3.1) | 103 (3.0) | 102 (2.6) | 651 (3.0) |
| BI-RADS 1 | 2,213 (15.6) | 469 (13.8) | 596 (15.1) | 3,278 (15.2) |
| BI-RADS 2 | 5,818 (41.0) | 1,416 (41.6) | 1,378 (35.0) | 8,612 (40.0) |
| BI-RADS 3 | 1,292 (9.1) | 331 (9.7) | 333 (8.5) | 1,956 (9.1) |
| BI-RADS 4 | 2,601 (18.3) | 667 (19.6) | 956 (24.3) | 4,224 (19.6) |
| BI-RADS 5 | 94 (0.7) | 27 (0.8) | 40 (1.0) | 161 (0.7) |
| BI-RADS 6 | 1,205 (8.5) | 298 (8.8) | 385 (9.8) | 1,888 (8.8) |
| **Background parenchymal enhancement** |  |  |  |  |
| Minimal/mild | 8,933 (62.9) | 2,117 (62.2) | 2,498 (63.5) | 13,548 (62.9) |
| Moderate/marked | 3,309 (23.3) | 858 (25.2) | 1,068 (27.1) | 5,235 (24.3) |
| **Fibroglandular tissue** |  |  |  |  |
| A/B (Fatty/scattered) | 4,664 (32.8) | 1,094 (32.1) | 1,304 (33.1) | 7,062 (32.8) |
| C/D (Heterogeneous/extreme) | 7,295 (51.4) | 1,796 (52.8) | 2,178 (55.3) | 11,269 (52.3) |

**Table 2:**
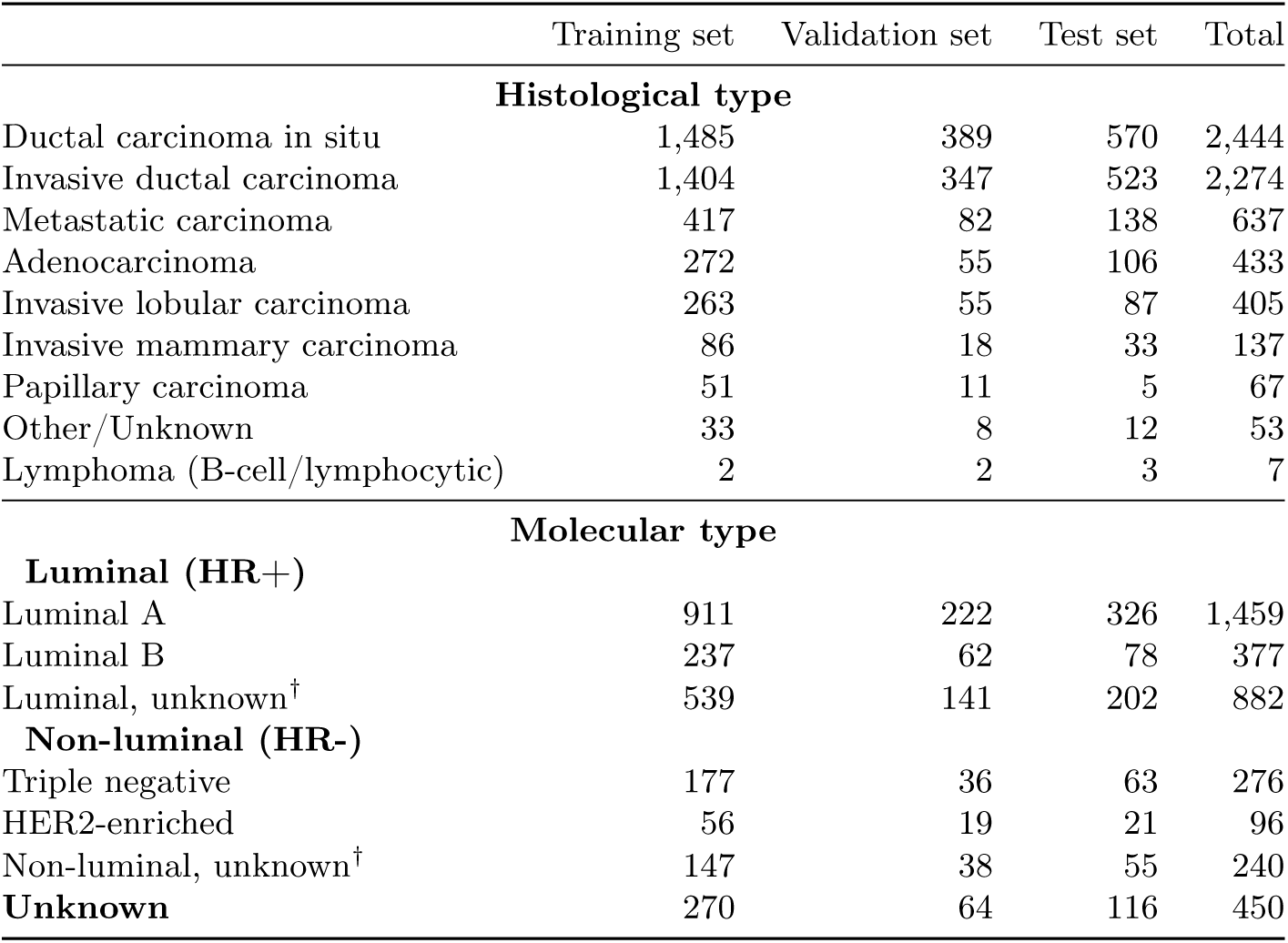
Histological and molecular cancer subtypes. One patient and one study can have multiple findings. †Because many studies in our data set had missing information about HER2 receptor status and/or Ki-67 status, they only have partial luminal/non-luminal classification. For example, if cancer was classified as ER+/PR+ but the information about HER-2 status was missing, it is considered “luminal, unknown”. Values are *N* (% of all malignant cases), reported on a study level. *HR*, hormone receptor; *ER*, estrogen receptor; *PR*, progesterone receptor; *HER2*, human epidermal growth factor receptor 2.

We validate our model performance on three international external data sets. The first data set was collected at the Jagiellonian University Hospital (Kraków, Poland) between 2019-2021 and contains 397 DCE-MRI studies. The remaining two data sets are as follows: 922 MRI studies from Duke University (18, 19) and 131 studies from the TCGA-BRCA data set (20). Characteristics and processing of all data sets are described in Methods, and the MRI scanner breakdown is in Table S3.

### Primary model performance

The AI system’s output is a prediction of the breast-level probability of malignancy. That is, for each of the patient’s breasts, the system produces a number in a range between 0 and 1. The AI system achieved 0.924 AUROC (95% CI: 0.915-0.933) and 0.720 AUPRC (0.689-0.751) when classifying breast cancer studies in the internal test set (Figure 2). Partial AUROC (pAUC) at 90% sensitivity was 0.765 (0.738-0.793) and 0.817 (0.801-0.833) at 90% specificity.

**Figure 1:**
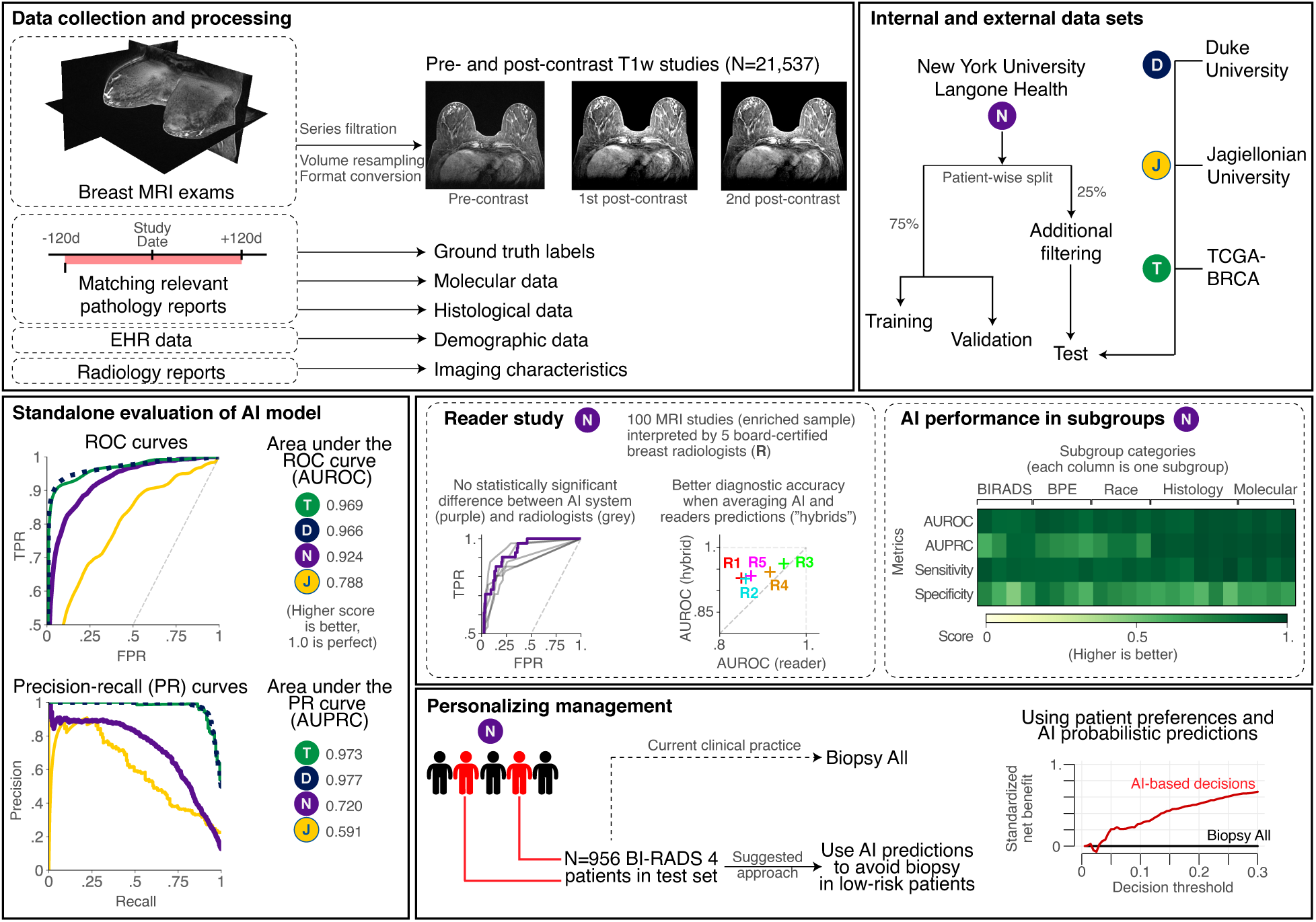
Overview of the study. In this work we train and evaluate an AI system based on deep neural networks that predict the probability of breast cancer in DCE-MRI studies. Our system takes one pre-contrast and two post-contrast T1-weighted fat saturated sequences as inputs and returns a probability of malignancy for each breast. We use a large NYU Langone data set for model training, validation and testing. Additionally, we obtained and processed three international data sets from Duke University (USA), Jagiellonian University (Poland) and TCGA-BRCA data set. We use these external data sets for the evaluation of system performance on out-of-distribution data. Besides traditional standalone evaluation, we look into performance in key patient subgroups, and we compare model accuracy to board-certified radiologists. Finally, we show that using predictions from our AI system can successfully personalize management of BI-RADS 4 patients and increase clinical utility by avoiding unnecessary biopsies.

**Figure 2:**
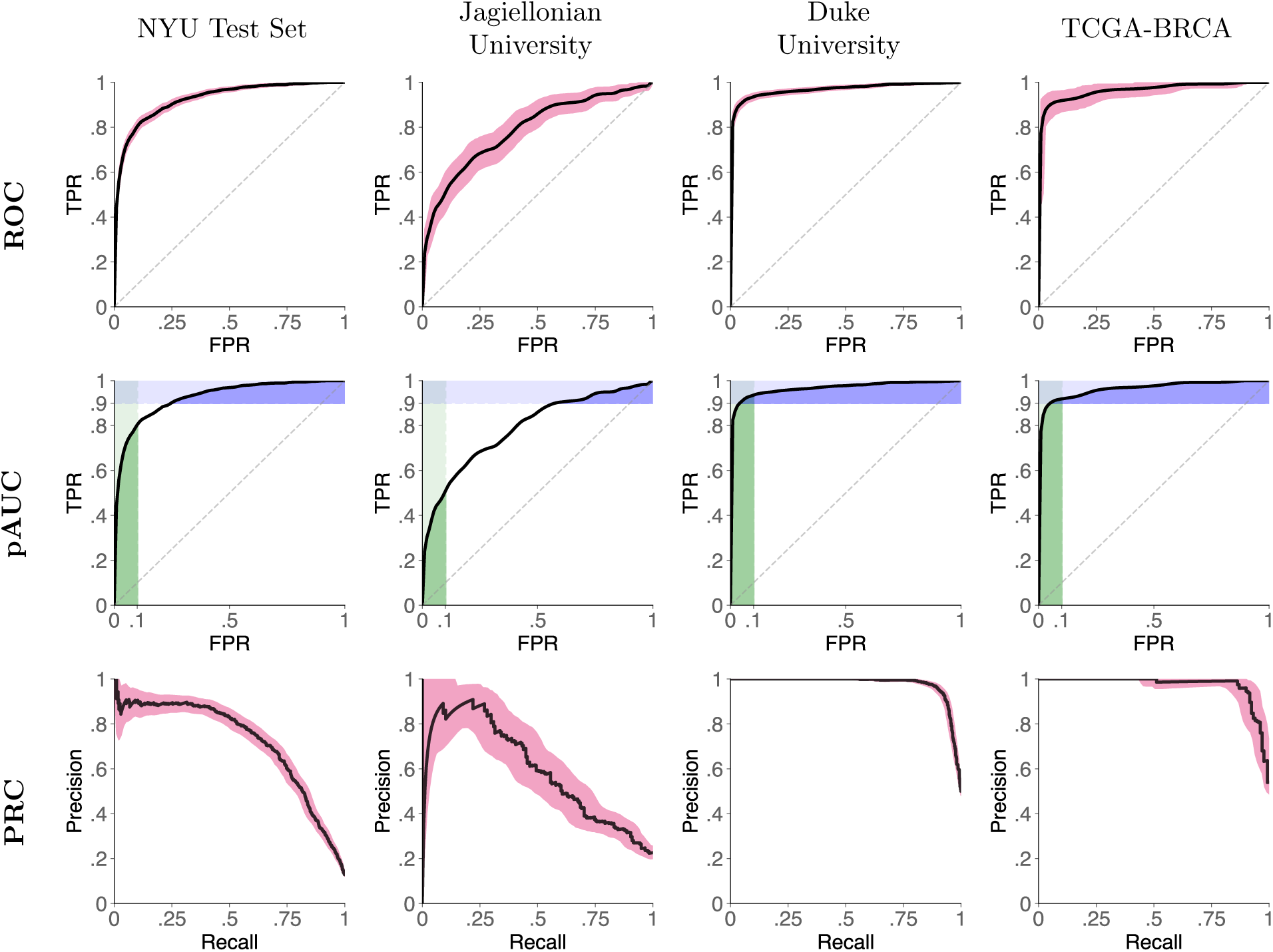
AI system performance on all test sets. Figures show receiver operator characteristic curves (ROC, top row), ROC curves with partial area under the curve (pAUC, bottom row), and precision-recall curves (PRC, bottom row) for all test sets. In the bottom row, green rectangles represent pAUC for 90-100% specificity and blue rectangles represent pAUC for 90-100% sensitivity. ROC and PR curves are presented with 95% confidence intervals calculated with bootstrapping (N=2,000).

On the Jagiellonian University test set, our system achieved 0.788 AUROC (0.750-0.832) and 0.591 AUPRC (0.514-0.672). On the Duke University data set, the AI system reached 0.969 AUROC (0.960-0.976) and 0.977 AUPRC (0.971-0.982), and on the TCGA-BRCA data set 0.966 AUROC (0.942-0.985), 0.973 AUPRC (0.954-0.988).

In our retrospective reader study, five (5) radiologists interpreted 100 cases sampled from the NYU Langone test set. All radiologists were board-certified and had between 2 and 12 years of experience interpreting breast MRI exams. Readers achieved an average performance of 0.890 AUROC (range: 0.850-0.948) and 0.758 AUPRC (range: 0.712-0.868). The AI model standalone performance on the reader study subset was 0.924 (0.880-0.962) AUROC and 0.784 (0.656-0.887) AUPRC. This was better than the average reader performance by 0.035, but the difference between AI and radiologists was not statistically significant (Obuchowski-Rockette model, 95% CI AUC difference: 0.09, −0.02; p=0.19). In a head-to-head comparison between the AI system’s and radiologists’ AUROC, the AI was significantly better than two of the readers (#1 and #2). ROC curves for all readers are presented in Fig. S1 and numerical results are in Table S1.

We evaluated the performance of *hybrid models* by averaging radiologists’ and AI system’s predictions. On average, an equally weighted hybrid improves AUROC by 0.05 and AUPRC by 0.07. We demonstrate benefits were seen in all readers when using averaging AI’s and radiologists’ predictions (Fig. S2). When changing weights of the hybrid, we found that using AI predictions, even with a very small weight (e.g. 5-10% weight), significantly improves the overall outcome. This weight could be treated as an operating point and be set specifically for different readers, as the optimal operating point (that maximizes the performance) varies between radiologists (Fig. S3).

### Subgroup analyses

We conducted analyses on a wide range of patient subgroups in the test set, dividing them with respect to demographic data, imaging features, and other characteristics of patients, studies and cancers. This was to establish if the model is biased in any of the evaluated categories and investigate if some groups are more likely to benefit more from the model. Results are presented in Figure 3, full numerical values are in Table S2 and select ROC/PR curves are in Figures S4 and S5.

**Figure 3:**
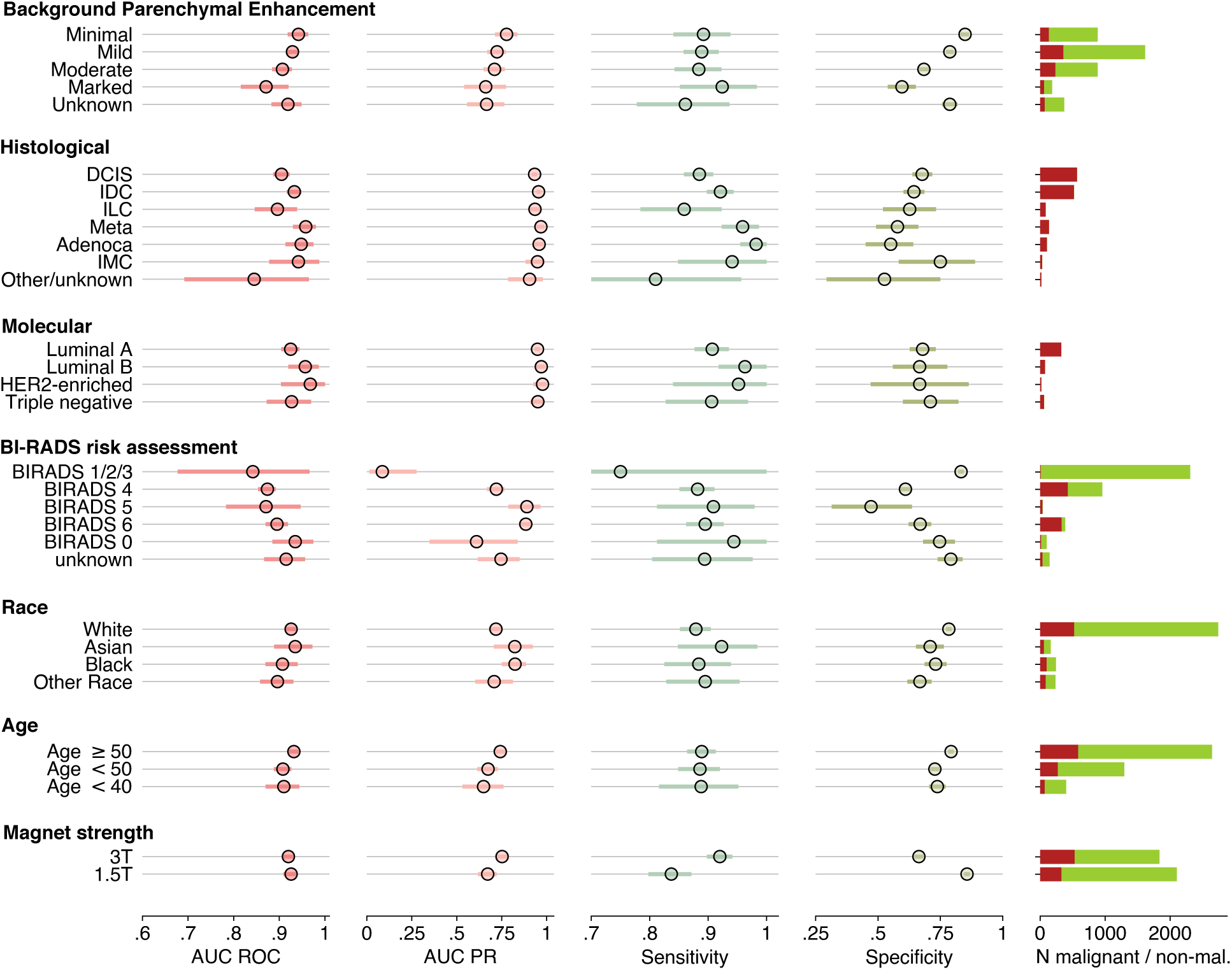
System performance in key subgroups on the internal test set. Plots present metric breast-level values with 95% confidence intervals. A decision threshold was selected such that the AI system’s sensitivity closely matches average reader sensitivity. CIs were calculated with bootstrap (N=2,000 replicates). Full numerical values for each subgroup are available in Table S2. BI-RADS categories 1, 2 and 3 are aggregated because there were no malignant cases in BI-RADS 1 and 2 categories, thus AUROC would not be defined in those subgroups. On the right, a number of malignant (red) and non-malignant (green) studies are presented.

**Figure 4:**
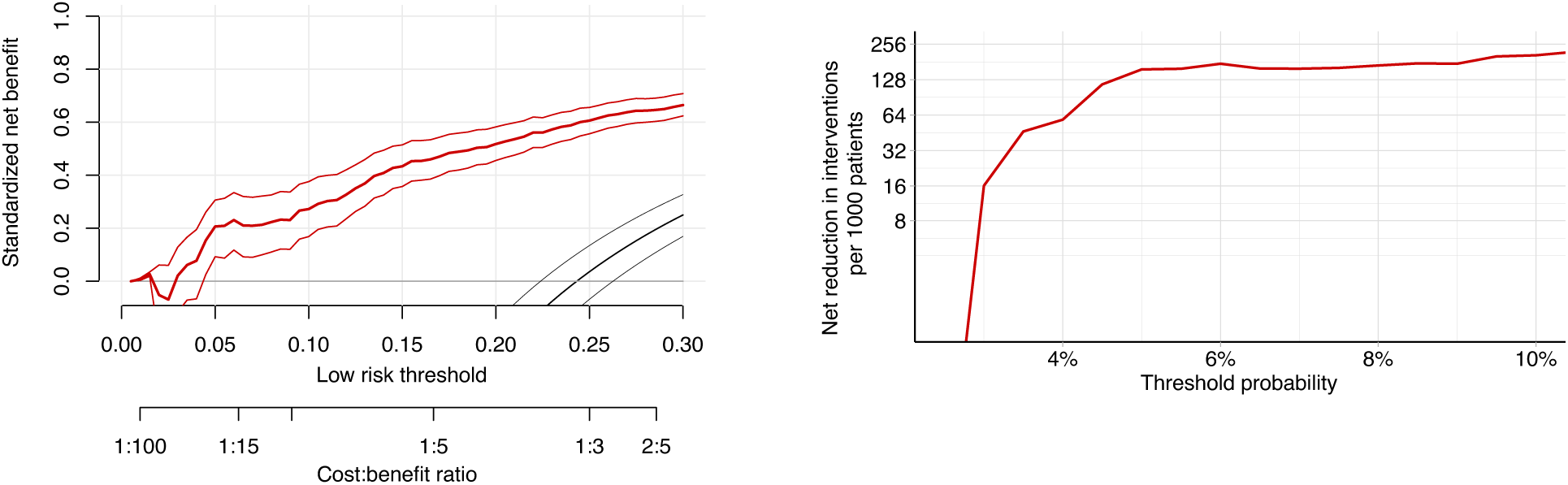
Decision curve analysis supports using the AI system to decide on whether to perform biopsy in low-risk BI-RADS 4 patients. **a**, standardized net benefit values are higher when decisions are made based on the AI system’s predictions (red curve) compared to default biopsy-all approach (grey line at *x* = 0) across all relevant decision thresholds (black curve is a biopsy-none approach). Net benefit curve is presented with 95% bootstrapped confidence intervals (N=2,000 replicates). **b**, shows net interventions avoided per 1,000 BI-RADS 4 patients. Reductions are highest when decision threshold is above 3%.

There were no noticable differences in the AI system’s performance between patients with various histological cancer subtypes, even when comparing more common cancers (e.g. invasive ductal carcinoma) with less common malignancies (e.g. invasive lobular carcinoma, ΔAUC: 1.5; two-sided DeLong’s test p=0.15). When considering patient demographics, the results indicate that the AI system appears to be unbiased, even when the subgroup was not as commonly represented in the training set, e.g. Black women (N=802 patients in the training set; test set AUROC 0.91 [0.87-0.95]) versus White women (N=9,819 in the training set; test set AUROC 0.93 [0.91-0.94]). AUC difference between those two groups was not statistically significant (p=0.33).

There is a slight trend of performance deteriorating in women with more significant background parenchymal enhancement (BPE). BPE is defined as the enhancement of the normal breast tissue in DCE-MRI, and often confounds the interpretation of this exam. The system was performing best in women with minimal BPE (AUROC 0.94 [0.92-0.96]) and gradually worsening in women with higher BPEs (marked: 0.87 [0.82-0.91]). AUC difference was statistically significant between minimal and marked BPE (ΔAUC: 2.5; p=0.01), mild and marked BPE (ΔAUC: 2.1; p=0.03) and minimal and moderate BPE (ΔAUC: 2.2; p=0.03).

### Personalized management of BI-RADS 3 and BI-RADS 4 patients

To create a diagnostic decision making application that allows for better concordance with patient preferences, we explore the possibility of using the AI model predictions as an aid in downgrading patients with BI-RADS category 4 to BI-RADS category 3, enabling them to opt out of a breast biopsy. This analysis is conducted in two ways, both using the full test set and original BI-RADS of the studies as reported initially by radiologists. First, we directly compare trade-offs between correctly avoided biopsies and missed cancers at various decision thresholds used to binarize probabilities of malignancy. This trade-off is an equally weighted comparison between the number of successfully opted-out patients (patients who avoided unnecessary biopsy) and missed cancers. In a second, more advanced approach, we use decision curve analysis methodology to incorporate patients’ and clinicians’ preferences into decision-making and explore whether using the model can be clinically beneficial or harmful.

At an operating point that leads to avoiding benign biopsies in 5.4% of all non-malignant BI-RADS 4 cases, no cancers would be missed. At a different operating point, if 22.9% non-malignant BI-RADS 4 patients avoided a biopsy, 10 (2.3% of all cancers in BI-RADS 4 cases) cancers would be missed. Increasing the threshold causes the model to successfully opt-out more patients, but also to miss more cancers (Figure S6). We investigated several cases that would be missed following this approach, and discovered that most missed cancers were ductal carcinomas in situ. They were usually described on the initial radiology report as “clumped nonmass enhancements”, sometimes associated with post-biopsy clips.

Similarly, patients with BI-RADS category 3 could potentially be downgraded to BI-RADS category 2, subsequently leading to their return to routine screening instead of short-term follow-up MRI after 6 or 12 months. As patients with BI-RADS category 3 rarely end up being diagnosed with a breast cancer, our model allows to correctly downgrade 235 BI-RADS 3 patients (73.2% of all non-malignant BI-RADS 3 cases) to BI-RADS 2, missing three cancer cases).

Using decision curve analysis (DCA) methodology (21), we evaluate clinical usefulness of our AI system at various decision thresholds. Unlike the technique described above (trade-off at various operating points), this approach is more informative as it takes into account the value of missed cancers versus the value of unnecessary interventions. The clinical decision we evaluated was whether the AI system’s predictions can select low-risk BI-RADS 4 patients and opt them out from undergoing a biopsy (effectively downgrading these patients to BI-RADS 3). Default practice is to biopsy all BI-RADS 4 patients, but this leads to an unreasonable number of false positive biopsies. With DCA we show that using AI’s predictions to opt-out low-risk patients is almost always better than performing biopsies for all patients.

Our results show that using the AI system in BI-RADS 4 patients offers net reduction in interventions even in low decision thresholds. If a decision threshold is at 5%, our approach offers a net reduction of 156 breast biopsies per 1,000 patients. In higher decision thresholds, benefits are even greater, up to approximately 200 net breast biopsies avoided per 1,000 patients at 10% threshold. We do not evaluate even higher decision thresholds, as we believe they are rarely clinically relevant (e.g. in older patients with very strong preferences) and do not reliably represent most patients’ or clinicians’ preferences. In sub-2% decision thresholds there is no net benefit in using our system, but these patients are not a subject of interest in this analysis and should always undergo a biopsy.

### Averaging predictions improves interreader variability

When considering positive versus negative studies (positive defined as assigned a probability of malignancy greater than 2% [i.e., a likelihood of malignancy that guides radiologists to rate a study as BI-RADS 4A]), mean agreement between readers was moderate for both breast-level (*κ*: 0.56 [95% CI: 0.50-0.63]) and exam-level (*κ*: 0.55 [0.46-0.64]) decisions. When averaged with AI predictions, interreader agreement improved to substantial, with a mean breast-level *κ* of 0.77 (0.72-0.82). The mean improvement in *κ* agreement was 0.21 (0.17-0.26). Similarly, intraclass correlation coefficient (ICC) improved from 0.663 (0.6-0.72) for readers only to 0.881 (0.85-0.90) for equally weighted hybrids between radiologists and AI.

### Reader study error analysis

Finally, we performed an error analysis of our system’s predictions using the reader study subset. We compared predictions of malignancy made by AI to predictions made by radiologists and investigated cases if there were major discrepancies. In a significant majority, AI system predictions matched those of radiologists. AI system behaved correctly when it predicted studies with cancer by giving them a high probability of malignancy, as well as when it correctly identified negative studies by giving them a very low probability. Moreover, in some benign cases AI’s predictions were much lower than radiologists, further demonstrating potential to avoid unnecessary biopsies. Having said that, we identified a few situations where our model gave unreasonably small probability of malignancy in cancer cases. We were not able to indicate a reason for this behavior, which shows a need for future research on AI uncertainty and explainability. Details of error analysis can be found in Supplementary Materials (section 9).

## DISCUSSION

In our study, we showed that our AI system reaches a very high standalone performance, equivalent to breast radiologists, and that it has a significant potential to personalize patient management, reducing the number of unnecessary biopsies in BI-RADS 4 patients.

Our study is the first to report performance on both internal and external MRI data sets (22). These data sets vary in terms of ground truth definition, as well as patient demographics, inclusion criteria, indication for the examination, acquisition protocols, and more. Because of this, drawing conclusions about the system’s performance “in-distribution” from the external data sets only may not be representative. On the other hand, this analysis should be good indication of the performance in data sampled from distributions not seen before by the model. For instance, our network had near-perfect performance on data sets that included only cancer cases (Duke University and TCGA-BRCA). On the other hand, our system yielded weaker results using the Jagiellonian University data set. We hypothesize that this difference is mostly a result of dissimilarity in either ground truth definition or data variability. Specifically, the Polish dataset includes all study indications, meanwhile our test set was heavily filtered (see Methods) to unify the patient population and reduce label noise.

A recent meta-analysis identified eight studies on the use of AI in MRI (22), all of them with very limited test sets. In that meta-analysis, an average AUC of included studies was 0.868 (95% CI: 0.850–0.886). While our study demonstrated that our AI system has higher AUC, a direct comparison to these studies is not possible as the exact definitions of the task and the test sets are different.

Previously, a few breast imaging studies explored the potential to downgrade BI-RADS 4 patients using predictive models, including AI-based algorithms. Works by Wang et al. (23), Zhao et al. (24) and Xiao et al. (25) assessed a commercially available AI tool for breast ultrasound to downgrade category 4A cases to category 3. In the Wang et al., out of 43 category 4A studies, 14 were correctly downgraded. In the Xiao et al. study, 23 out of 42 category 4A lesions (40 benign and 2 malignant) were correctly downgraded.

Our study also evaluates clinical utility by following a decision curve analysis (DCA) methodology. DCA indirectly incorporates patient and doctors preferences and calculates a “net benefit” of a new strategy (e.g. only biopsying patients who have high AI-based risk) compared to the default (e.g. biopsy all). DCA does not explicitly ask doctors/patients about their preferences, but rather calculates the net benefit across a range of threshold probabilities. These probabilities represent the trade-off of preferences, considering concerns about both cancer and biopsy. While DCA is a relatively new methodology, it has been encouraged by medical journals and adopted in several publications (26–28). In our analysis, we saw benefits in using the AI system to avoid unnecessary biopsies compared to performing them in all BI-RADS 4 patients. These benefits were consistent in all clinically relevant threshold probability values. We encourage the readers to refer to Vickers et al. for a thorough analysis on how to interpret DCA results (29).

We acknowledge a few limitations of this study. First of all, our reader study design and averaging radiologists predictions with AI system outputs might not capture how the system will affect radiologists decision-making, once implemented in hospital systems. A multireader, multicase study, in which an AI system would be used as a concurrent or second read tool that complies with regulatory guidelines, would be of value. Second, our decision curve analysis gives valuable information about clinical utility, but in the future a more extensive, cost-effective analysis should be performed to estimate monetary benefits and acceptable costs of the AI system. Third, we did not investigate whether our system learned to use minor clues in images, such as a biopsy clip marker artifact, to affect its decisions. Finally, our system does not provide clear explanations of its decision making. Even though we make the process of AI development transparent, predictions given by the system do not point to a specific part of the image or patient characteristic that influenced the prediction.

In conclusion, our model achieves a level of diagnostic accuracy equivalent to breast imaging experts for predicting the presence of breast cancer in DCE-MRI studies. We showed this high performance generalizes well to other populations through evaluation on international, external data sets. Combining AI system’s predictions with radiologists decisions further improves classification accuracy. Clinically, our model can help personalize patient management, leading to a reduced number of unnecessary workup and biopsies, and is better than a biopsy-all strategy in BI-RADS 4 patients.

## MATERIALS AND METHODS

### Study Design

#### Ethical approval

This is a retrospective study that was approved by the Institutional Review Board (ID #s18-00712). Informed consent requirement was waived. The study follows the guidelines of the Declaration of Helsinki of 1975 with its later revisions.

#### NYU Langone Health data set

The data set consists of 21,537 DCE-MRI studies from 13,463 patients who underwent DCE-MRI at NYU Langone Health between 2008 and 2020. The data was randomly split into training, validation and test sets with 60%, 15% and 25% of the data, respectively. This split was made on a patient-level, so that data from one patient can be in only one subset.

#### Ground truth

All data have breast-level labels that describe the presence or absence of benign or malignant findings in either the left and right breast. All benign and malignant labels in our data set are pathology-proven, based on specimen analysis from either a breast biopsy or surgery. We searched and matched all pathology reports dated 120 days before or after the day of examination.

#### Filtering the data set

To maximize the accuracy of our truthing, and to remove potentially confounding subgroups, we performed additional filtering of our data set. In the full data set, we excluded studies where we were not able to reliably determine a cancer label or there were technical issues with data extraction or data consistency. We also added a set of rules specific to the test set to further remove the label noise. We excluded patients with a history of bilateral mastectomy (N=165) and patients after neoadjuvant chemotherapy (N=105). We do not intend to use our software in those sub-populations, though we encourage future researchers to take this into consideration. Additionally, we excluded patients with breast implants (N=370). We also manually reviewed cases where: *(1)* a study was initially labeled as negative, but was assigned BI-RADS 1, 2 or 3; *(2)* a study was labeled as malignant, but was assigned BI-RADS 1 or 2; *(3)* a study was labeled as malignant, but was assigned BI-RADS 0, 3 or 6. In situation *(1)*, we added a 1 year negative follow-up requirement. Negative follow-up means that in a year after the MRI study date: *(1)* there are no pathology reports associated with the patient; *(2)* there is at least one breast imaging study (mammography, MRI) with BI-RADS category 1, 2 or 3; *(3)* there are no breast imaging studies with BI-RADS category 0, 4, 5 or 6. In situation *(2)*, we excluded those cases as it is a clear mistake to label a study as both malignant and BI-RADS 1 or 2. In situation *(3)*, we manually reviewed all cases (N=433) and verified correctness of the labels. If necessary, studies had their labels fixed or were excluded. A flowchart with details about overall data set and test set specific filtering, please refer to our data report (30).

#### External data sets

To prove generalizability of our model in a different population, we collected data sets from multiple institutions in the United States and Poland. All external data underwent the same preprocessing pipeline as the NYU Langone data set. That is, they were resampled, reoriented to the LPS (left-posterior-superior) orientation and saved in an appropriate file format.

#### Jagiellonian University (“JU Hospital”)

This data set has been acquired at the Jagiellonian University Hospital, which is the highest-volume hospital in southern Poland. Breast cancer diagnosis and treatment is overseen by the JU Hospital Breast Unit, which is an accredited, multidisciplinary team specialized in breast cancer. Images in the JU Hospital data set were anonymized on by JU Hospital before being transferred to NYU Langone Health.

Labeling and anonymization was performed by a board-certified breast radiologist, and the labels are pathology-proven, similar to our internal data set. 99% of studies have been acquired on a 1.5T Siemens MAGNETOM Sola between December 2019 and August 2021. Indications for a scan varied, but the largest group was patients undergoing a follow-up after ambiguous findings in other modalities.

After obtaining the data set, we semi-automatically identified pre- and post-contrast sequences in all studies and converted them to the NIfTI format. Then, we performed a manual visual review of saved images to confirm accuracy of pre-/post-contrast assignment and found any studies that did not have fat-saturated series. From the original data set, we excluded 5 studies that did not contain fat-saturated images, 3 unilateral studies, and one study that did not have consistent image size in pre- and post-contrast series. Ultimately, 397 studies were included with 248 benign and 149 malignant studies.

#### Duke University

The Duke Breast Cancer MRI data set (“Duke data set”) is publicly available through The Cancer Imaging Archive (TCIA). It contains 922 studies (N=922 patients) with invasive breast cancer, scanned on either 1.5T or 3T machines. Authors of this data set also provide detection labels, clinical features and imaging features. Detection labels are available for all studies and are shared in a tabular form describing coordinates of 3D cuboid bounding boxes. Images in the Duke data set are stored in the DICOM format and were preprocessed with the same pipeline as NYU Langone data set. That is, they were resampled and reoriented to the LPS orientation. We identified pre- and post-contrast sequences, as required by our model, and performed inference to generate predictions. Because our model makes breast-level predictions, we converted bounding box labels to breast-level labels. New labels were produced based on the middle point of bounding boxes. For example, if the central point of a bounding box is located on the left anatomical side of the patient, a label for left breast is generated.

As described in Saha et al. (19), the images were annotated by eight fellowship-trained breast radiologists with 1-22 years of post-fellowship experience.

We excluded the following studies from the Duke data set: `Breast_MRI_065, Breast_MRI_120, Breast_MRI_127, Breast_MRI_134, Breast_MRI_232, Breast_MRI_279, Breast_MRI_465, Breast_MRI_514, Breast_MRI_574, Breast_MRI_596, Breast_MRI_700` and `Breast_MRI_767`. The reason for the exclusion was either size mismatch between pre- and post-contrast sequences, or an inability to automatically determine which sequences were pre- or post-contrast. Ultimately, 910 studies were included from the Duke data set. As the Duke data set is all-malignant, interpretation of its results should be limited to patients with invasive breast cancer.

#### The Cancer Genome Atlas Breast Invasive Carcinoma (TCGA-BRCA)

This is a data set publicly available as a part of The Cancer Imaging Archive (TCIA) (31), similar to the Duke University data set. Originally, this data set included 164 studies from 139 patients and contains images, clinical data, biomedical data and DICOM Structured Reporting files. The data in its original form is not suitable for AI evaluation, which is why we developed a pipeline for generating AI-ready TCGA-BRCA data with labels (Supplementary Materials 5). After processing, the data set contains 131 studies.

### AI system

#### Inputs and outputs

Let **x** ∈ ℝ^*C,Z,X,Y*^ denote an input. **Z, X, Y** are the spatial dimensions of a MRI volume, and **C** channels are different MRI sequences, i.e. pre- and post-contrast series. The neural network generates four probability estimates: *ŷ*_*lb*_, *ŷ*_*lm*_, *ŷ*_*rb*_, *ŷ*_*rm*_ [0, 1] that indicate the predicted probability of the presence of benign and malignant lesions in each of the patient’s breast (i.e. *b* and *m* represent benign and malignant findings, and *l* and *r* represent left and right breasts, respectively.). Probabilities of benign findings (*ŷ*_*lb*_, *ŷ*_*rb*_) are used only as a multi-task learning regularization method. Throughout the study we only evaluate predicted probabilities of malignant findings (*ŷ*_*lm*_, *ŷ*_*rm*_).

#### Architecture

On a high level, our models are deep residual neural networks with 3D convolutions to detect spatiotemporal features (32). Specifically, we use a 3D-ResNet18 backbone with a max pooling layer before linear classifier.

In our experiments, we found that models with pre-trained backbones perform better compared to models trained from scratch. We used weights from models pre-trained on the Kinetics-400 data set, which is an action recognition in videos data set (33). Even though Kinetics-400 is not a medical data set, transfer learning was successful to our downstream task. This is in line with other results suggesting that medical imaging can benefit from large-scale non-medical data sets for transfer learning (34, 35).

#### Data augmentation during training

We experimented with various geometrical, intensity-based and MRI-specific augmentations (36). Ultimately, in our experiments we found that affine transformation gave the most consistently beneficial results. For all models, during training, we randomly (p=0.5) flipped images “horizontally”, i.e. in the left-right anatomical axis. Note that when the volume was flipped, labels were flipped as well, i.e. *y*_*lb*_ ← *y*_*rb*_ while *y*_*rb*_ ← *y*_*lb*_ and *y*_*lm*_ ← *y*_*rm*_ while *y*_*rm*_ ← *y*_*lm*_.

Note that the affine augmentations applied were not based on the tensor sizes, but rather on the real-life dimensions. To convert into anatomical dimensions, we calculated affine matrices for each study that define relationships between pixel and real life sizes. To compute an affine matrix, we used image spacing, origin and direction cosines values that were collected from DICOM metadata. Those matrices were necessary to initially resample to the same pixel spacing and reorient all images to the LPS (left-posterior-superior) orientation, which is a standard orientation in DICOM convention. That means that pixels in the matrix follow the anatomical order and go from right towards left, from anterior towards posterior and from inferior towards superior. Resampling was performed using linear interpolation.

#### Subtraction images

During our experiments, we attempted to improve the performance by using subtraction images. They were generated by a simple matrix subtraction between post-contrast and pre-contrast volumes, such that 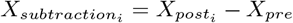, where *X*_*post*_ is one of *i* post-contrast volumes and *X*_*pre*_ is the pre-contrast volume. We did not clip lower range values to zero, which is sometimes done by radiological software to improve visibility. Ultimately, we did not find subtraction images provided superior performance over regular T1-weighted sequences. We hypothesize that the neural network, when provided with pre- and post-contrast images, is able to learn patterns related to the contrast enhancement.

#### Training details

In our study, reported performance for the “best model” is using an ensemble of top 20 models. All models were trained with the Adam optimizer (37), and top models for the ensemble were selected after hyperparameter tuning with random search (38), as described below. AUROC for malignant labels was the target metric in the hyperparameter search. We tuned the following parameters:

- scaling ∈ [5%, 25%], symmetrically along each axis,
- rotation ∈ [5 deg, 30 deg], symmetrically along each axis,
- translation ∈ [5 mm, 20 mm], symmetrically along each axis,
- dropout - 25% chance on the fully connected layer (yes or no),
- label smoothing (39, 40) *α* ∈ [0, 0.1],
- stochastic depth rate (41) ∈ [0, 0.1],
- weight decay ∈ [1*e*−6, 1*e*−4],
- learning rate ∈ [7*e*−7, 2*e*−5],
- number of warmup epochs ∈ [2, 6],
- choice of the learning rate scheduler (10x reduction after *α* epochs versus cosine annealing) and scheduler policies.

Models were trained with mixed precision using NVIDIA Apex open source library. Network architecture included group normalization (42) instead of batch normalization (43), as large volume size dramatically reduces the number of samples in a mini-batch. We found that group normalization with 16 groups was performing best in our experiments. We used Neptune.ai and Weights&Biases for tracking, evaluating and visualizing experimental results (44, 45).

#### Test time augmentations

During test time, we transformed all data samples 10 times (test time augmentation, TTA), and averaged inference results from all TTA samples. This approach has been shown to improve accuracy and robustness of AI models. The optimal TTA policy was found on a validation set after running a random search for TTA hyperparameters: number of TTA rounds ∈ [1, 10], affine scaling ∈ [10%, 20%], rotation ∈ [10 deg, 20 deg], translation [10 mm, 20 mm], using gamma transformations or blurring. On our validation set, we found that the best performing TTA policy was the one that implemented 10 rounds of TTA, random horizontal flips, and affine transformations of 10% scaling factor, 10 degree rotation and 10 pixel translation. However, we also note that differences between various TTA policies were usually indistinguishable. For single models (i.e. not full ensemble), we noticed an improvement of ≈0.005 AUC ROC and ≈0.01 AUC PR on our validation set.

#### Reader study

We designed a retrospective reader study to compare the standalone clinical performance of our model with radiologists. This study had a single-arm design (readers interpreting MRI studies only). We recruited 5 board-certified breast radiology attendings to participate in the study. We randomly selected 100 studies from the primary test set as a *reader study set* for readers to interpret. Following the common practice in reader studies, our data set has been enriched with malignant and non-malignant biopsied cases. Specifically, there were 40 malignant, 40 benign, and 20 negative studies in the reader study set. Readers were informed that the population of the study does not represent typical distribution of patients undergoing breast MRI, but they had no knowledge about the specific split. Readers were also blinded to any confidential information, prior studies or indications for the examination. Radiologists had access to all available MRI sequences, and were not limited to T1-weighted fat-saturated series that are used as inputs for the AI system.

Readers were provided with a workstation preloaded with studies. All studies used in the reader study have been pseudonymized and stored in a DICOMweb server (46) ^1^ separate from clinical PACS servers. Radiologists had access to the workstation and a data collection tool where they reported their predictions. Before joining the study, recruited readers had to familiarize themselves with study instructions as well as the DICOM viewer (47) used in the study. Radiologists had to provide the following predictions:

- A probability of malignancy (POM) for the whole study, in a range between 0 and 100.
- A POM for the left breast, in a range between 0 and 100.
- A POM for the right breast, in a range between 0 and 100.
- A “forced BI-RADS” for the study. This means they could only use BI-RADS categories 1, 2, 3, 4A, 4B, 4C or 5. BI-RADS categories 0 or 6 were not available. If the study was classified as BI-RADS 4, radiologists had to specify a subcategory (4A, 4B or 4C).

Readers were advised to use BI-RADS likelihood of cancer ranges as a guideline when assigning POM values. This means that if a reader believes that the study is probably benign (BI-RADS 3), they should assign a POM value in the (0, 2] interval, etc.

#### Statistical analysis

To measure model performance, we used receiver operating characteristic (ROC) and precision-recall (PR) and calculated areas under the curve (AUC) for both (AUC ROC and AUC PR). We also report sensitivity and specificity. Areas under curves were calculated with a non-parametic (trapezoidal) method. We further evaluate specific clinical scenarios using partial AUC (48, 49) statistic, as implemented in the pROC R package (50). All results, where appropriate, are reported with 95% confidence intervals using bootstrapping (51). When evaluating standalone AI performance versus reader performance, we used a single-treatment random-reader random-case model, based on the Obuchowski-Rockette model (52, 53). This method takes into account variability both between readers, and between cases. We used the model based on descriptions by Chakraborty (54) and implementation from the *RJafroc* (55) R package (the “1T-RRRC” model). Null hypothesis for significance testing was that the average breast-level AUC of the AI model equals the average AUC of radiologists. In subgroup analyses, for comparisons of AUCs of two curves, we performed a two-sided DeLong’s test (56).

For decision curve analysis, we used R packages *rmda* and *dcurves* to generate the curves, calculate net benefit and net interventions avoided. We established a range of reasonable threshold probabilities for evaluated subgroups (BI-RADS 4 and 3), and also reported a wide range of threshold probabilities on a full population.

The standardized net benefit (sNB; a.k.a. relative utility) for the opt-out policy (here: downgrading BI-RADS 4 to BI-RADS 3) can be defined as:

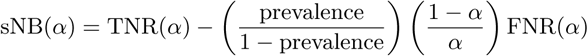

where prevalence is disease prevalence, *α* is decision threshold, TNR is a true negative rate and FNR is a false negative rate.

To avoid overestimating the net benefit (57), we bootstrapped the results with N=2,000 replicates and reported decision curves with 95% confidence intervals, as suggested by Kerr et al. (58).

For measuring interreader variability, we used Fleiss’ kappa (59), specifically with Randolph’s free-marginal modification (60) for agreement between positive and negative cases. We also used intraclass correlation coefficient (ICC) for measuring consistency in scoring probability of malignancy. Both Fleiss’ kappa and ICC were calculated on exam-level and breast-level for readers. ICC estimates are based on single measures with a two-way random-effects model, i.e. ICC(2,1) (61).

## Data Availability

Our internal (NYU Langone Health) data set is not publicly available due to internal data transfer policies. We released a data report on data curation and preprocessing to encourage reproducibility. The data report can be accessed at https://cs.nyu.edu/~kgeras/reports/MRI_datav1.0.pdf. The Jagiellonian University data set has been acquired for NYU Langone Health researchers through a data transfer agreement, and is not publicly available. Duke Breast Cancer MRI and TCGA-BRCA are publicly available in the Cancer Imaging Archive.

## Supplementary Materials

1. Reader study results. Figure S1. All receiver operating characteristic (ROC) and precision-recall (PR) curves from the reader study. Table S1. Reader study results.
2. Hybrid predictions. Table S1. Reader study results. Figure S2. Hybrid predictions are stronger than readers’ predictions alone. Figure S3. Performance of a hybrid model, as a function of *α* ∈ (0, 99]%.
3. Subgroup performance. Table S2. Subgroup performance. Figure S4. Empirical ROC curves for subgroups. Figure S5. Empirical precision-recall curves for subgroups.
4. BI-RADS downgrading. Figure S6. Trade-off in missed cancers versus correctly avoided interventions when using only AI system to decide on management.
5. Processing TCGA-BRCA data set.
6. Manufacturers and devices. Table S3. MRI manufacturer and model breakdown for all data sets.
7. Breast-level labels. Table S4. Breast-level breakdown in the NYU Langone data set.
8. Distribution of predicted probabilities of malignancy. Figure S7. Distribution of predicted probabilities of malignancy (POM) on the NYU Langone test set.
9. Error analysis. Figures S8-S17. Error analysis.

## Acknowledgements

This work was supported in part by grants from the National Institutes of Health (P41EB017183, R21CA225175) and the Gordon and Betty Moore Foundation (9683). JW was supported in part by The Iwanowska Programme from the Polish National Agency for Academic Exchange (PPN/IWA/2019/1/00114/U/00001). The authors would like to thank Mario Videna, Abdul Khaja and Michael Costantino for supporting our computing environment.

## Authors contributions

KJG conceived the idea for this study. JW and KJG designed experiments with neural networks. JW built the data preprocessing pipeline and performed experiments and result synthesis. LH, BR, AL, KP, SP, NS, LM collected the data. LH, BR, SK, AL analyzed the results from a clinical perspective. SK helped plan and analyze decision curve analysis results. WR, EŁ and TP assisted with Jagiellonian University data set acquisition and processing. KJG supervised the project. All authors contributed to drafting and review of the paper. All authors agreed to the final version of this manuscript.

## Data availability

Our internal (NYU Langone Health) data set is not publicly available due to internal data transfer policies. We released a data report on data curation and preprocessing to encourage reproducibility. The data report can be accessed at https://cs.nyu.edu/~kgeras/reports/MRI_datav1.0.pdf (30). The Jagiellonian University data set has been acquired for NYU Langone Health researchers through a data transfer agreement, and is not publicly available. Duke Breast Cancer MRI and TCGA-BRCA are publicly available in the Cancer Imaging Archive (18, 20). Because TCGA-BRCA data set originally does not provide information on pre- and post-contrast sequences as well as labels formatted for AI research, we publish new files and scripts that enable processing of the TCGA-BRCA data set for AI purposes. They are available at https://github.com/nyukat/MRI_AI.

## Code availability

Upon reasonable request, custom scripts used to process and analyze the data and generate figures will be made available by the authors of the manuscript. Authors will also consider requests for model weights and inference code. Requests are handled on a case-by-case basis depending on the nature of the request.

## Footnotes

1 https://github.com/dcmjs-org/dicomweb-server

## References

[1] R. M. Mann, C. K. Kuhl, and L. Moy, “Contrast-enhanced MRI for breast cancer screening,” Journal of Magnetic Resonance Imaging, vol. 50, no. 2, pp. 377–390, 2019.

[2] A.-j. Carin, S. Molière, V. Gabriele, M. Lodi, N. Thiébaut, K. Neuberger, and C. Mathelin, “Relevance of breast MRI in determining the size and focality of invasive breast cancer treated by mastectomy: a prospective study,” World Journal of Surgical Oncology, vol. 15, no. 1, pp. 1–8, 2017.

[3] C. D. Lehman, J. M. Lee, W. B. DeMartini, D. S. Hippe, M. H. Rendi, G. Kalish, P. Porter, J. Gralow, and S. C. Partridge, “Screening MRI in women with a personal history of breast cancer,” Journal of the National Cancer Institute, vol. 108, no. 3, p. djv349, 2016.

[4] C. K. Kuhl, K. Strobel, H. Bieling, C. Leutner, H. H. Schild, and S. Schrading, “Supplemental breast MR imaging screening of women with average risk of breast cancer,” Radiology, vol. 283, no. 2, pp. 361–370, 2017.

[5] A. R. Park, E. Y. Chae, J. H. Cha, H. J. Shin, W. J. Choi, and H. H. Kim, “Preoperative Breast MRI in Women 35 Years of Age and Younger with Breast Cancer: Benefits in Surgical Outcomes by Using Propensity Score Analysis,” Radiology, p. 204124, 2021.

[6] C. Spick, D. H. Szolar, K. W. Preidler, M. Tillich, P. Reittner, and P. A. Baltzer, “Breast MRI used as a problem-solving tool reliably excludes malignancy,” European Journal of Radiology, vol. 84, no. 1, pp. 61–64, 2015.

[7] B. Reig, “Radiomics and deep learning methods in expanding the use of screening breast MRI,” European Radiology, pp. 1–3, 2021.

[8] W. Lotter, A. R. Diab, B. Haslam, J. G. Kim, G. Grisot, E. Wu, K. Wu, J. O. Onieva, Y. Boyer, J. L. Boxerman, et al., “Robust breast cancer detection in mammography and digital breast tomosynthesis using an annotation-efficient deep learning approach,” Nature Medicine, vol. 27, no. 2, pp. 244–249, 2021.

[9] S. M. McKinney, M. Sieniek, V. Godbole, J. Godwin, N. Antropova, H. Ashrafian, T. Back, M. Chesus, G. S. Corrado, A. Darzi, et al., “International evaluation of an AI system for breast cancer screening,” Nature, vol. 577, no. 7788, pp. 89–94, 2020.

[10] X. Qian, J. Pei, H. Zheng, X. Xie, L. Yan, H. Zhang, C. Han, X. Gao, H. Zhang, W. Zheng, et al., “Prospective assessment of breast cancer risk from multimodal multiview ultrasound images via clinically applicable deep learning,” Nature Biomedical Engineering, vol. 5, no. 6, pp. 522–532, 2021.

[11] N. Wu, J. Phang, J. Park, Y. Shen, Z. Huang, M. Zorin, S. Jastrzębski, T. Févry, J. Katsnelson, E. Kim, et al., “Deep neural networks improve radiologists’ performance in breast cancer screening,” IEEE transactions on medical imaging, vol. 39, no. 4, pp. 1184–1194, 2019.

[12] Y. Shen, F. E. Shamout, J. R. Oliver, J. Witowski, K. Kannan, J. Park, N. Wu, C. Huddleston, S. Wolfson, A. Millet, R. Ehrenpreis, D. Awal, C. Tyma, N. Samreen, Y. Gao, C. Chhor, S. Gandhi, C. Lee, S. Kumari-Subaiya, C. Leonard, R. Mohammed, C. Moczulski, J. Altabet, J. Babb, A. Lewin, B. Reig, L. Moy, L. Heacock, and K. J. Geras, “Artificial Intelligence System Reduces False-Positive Findings in the Interpretation of Breast Ultrasound Exams,” medRxiv, 2021.

[13] A. Rodriguez-Ruiz, K. Lång, A. Gubern-Merida, M. Broeders, G. Gennaro, P. Clauser, T. H. Helbich, M. Chevalier, T. Tan, T. Mertelmeier, et al., “ Stand-alone artificial intelligence for breast cancer detection in mammography: comparison with 101 radiologists,” JNCI: Journal of the National Cancer Institute, vol. 111, no. 9, pp. 916–922, 2019.

[14] H.-E. Kim, H. H. Kim, B.-K. Han, K. H. Kim, K. Han, H. Nam, E. H. Lee, and E.-K. Kim, “Changes in cancer detection and false-positive recall in mammography using artificial intelligence: a retrospective, multireader study,” The Lancet Digital Health, vol. 2, no. 3, pp. e138–e148, 2020.

[15] S. L. van Winkel, A. Rodríguez-Ruiz, L. Appelman, A. Gubern-Mérida, N. Karssemeijer, J. Teuwen, A. J. Wanders, I. Sechopoulos, and R. M. Mann, “Impact of artificial intelligence support on accuracy and reading time in breast tomosynthesis image interpretation: a multi-reader multi-case study,” European Radiology, pp. 1–10, 2021.

[16] E. Verburg, C. H. van Gils, B. H. M. van der Velden, M. F. Bakker, R. M. Pijnappel, W. B. Veldhuis, and K. G. A. Gilhuijs, “Deep Learning for Automated Triaging of 4581 Breast MRI Examinations from the DENSE Trial,” Radiology, 2021.

[17] American College of Radiology, ACR BI-RADS Atlas: Breast Imaging Reporting and Data System. 5 ed., 2013.

[18] A. Saha, M. R. Harowicz, L. J. Grimm, J. Weng, E. H. Cain, C. E. Kim, S. V. Ghate, R. Walsh, and M. A. Mazurowski, “Dynamic contrast-enhanced magnetic resonance images of breast cancer patients with tumor locations [Data set],” 2021.

[19] Saha, M. R. Harowicz, L. J. Grimm, C. E. Kim, S. V. Ghate, R. Walsh, and M. A. Mazurowski, “A machine learning approach to radiogenomics of breast cancer: a study of 922 subjects and 529 DCE-MRI features,” British journal of cancer, vol. 119, no. 4, pp. 508–516, 2018.

[20] W. Lingle, B. Erickson, M. Zuley, R. Jarosz, E. Bonaccio, J. Filippini, and N. Gruszauskas, “Radiology Data from The Cancer Genome Atlas Breast Invasive Carcinoma [TCGA-BRCA] collection,” The Cancer Imaging Archive, vol. 10, p. K9, 2016.

[21] A. J. Vickers and E. B. Elkin, “Decision curve analysis: a novel method for evaluating prediction models,” Medical Decision Making, vol. 26, no. 6, pp. 565–574, 2006.

[22] R. Aggarwal, V. Sounderajah, G. Martin, D. S. Ting, A. Karthikesalingam, D. King, H. Ashrafian, and A. Darzi, “Diagnostic accuracy of deep learning in medical imaging: a systematic review and meta-analysis,” npj Digital Medicine, vol. 4, no. 1, pp. 1–23, 2021.

[23] X.-Y. Wang, L.-G. Cui, J. Feng, and W. Chen, “Artificial intelligence for breast ultrasound: An adjunct tool to reduce excessive lesion biopsy,” European Journal of Radiology, vol. 138, p. 109624, 2021.

[24] Zhao, M. Xiao, H. Liu, M. Wang, H. Wang, J. Zhang, Y. Jiang, and Q. Zhu, “Reducing the number of unnecessary biopsies of US-BI-RADS 4a lesions through a deep learning method for residents-in-training: a cross-sectional study,” BMJ open, vol. 10, no. 6, p. e035757, 2020.

[25] M. Xiao, C. Zhao, J. Li, J. Zhang, H. Liu, M. Wang, Y. Ouyang, Y. Zhang, Y. Jiang, and Q. Zhu, “Diagnostic value of breast lesions between deep learning-based computer-aided diagnosis system and experienced radiologists: comparison the performance between symptomatic and asymptomatic patients,” Frontiers in Oncology, vol. 10, 2020.

[26] M. Fitzgerald, B. R. Savilele, and R. J. Lewis, “Decision Curve Analysis,” JAMA, 2015.

[27] K. F. Kerr, M. D. Brown, K. Zhu, and H. Janes, “Assessing the Clinical Impact of Risk Prediction Models With Decision Curves: Guidance for Correct Interpretation and Appropriate Use,” Journal of Clinical Oncology, 2016.

[28] Deniffel, N. Abraham, K. Namdar, X. Dong, E. Salinas, L. Milot, F. Khalvati, and M. A. Haider, “Using decision curve analysis to benchmark performance of a magnetic resonance imaging–based deep learning model for prostate cancer risk assessment,” European Radiology, 2020.

[29] A. J. Vickers, B. van Calster, and E. W. Steyerberg, “A simple, step-by-step guide to interpreting decision curve analysis,” Diagnostic and Prognostic Research, 2019.

[30] J. Witowski, S. Gong, N. Wu, L. Moy, L. Heacock, B. Reig, S. G. Kim, F. Knoll, and K. J. Geras, “ The NYU Breast MRI Dataset,” tech. rep., 2021. Available at https://cs.nyu.edu/~kgeras/reports/MRI_datav1.0.pdf.

[31] K. Clark, B. Vendt, K. Smith, J. Freymann, J. Kirby, P. Koppel, S. Moore, S. Phillips, D. Maffitt, M. Pringle, et al., “The Cancer Imaging Archive (TCIA): maintaining and operating a public information repository,” Journal of Digital Imaging, vol. 26, no. 6, pp. 1045–1057, 2013.

[32] Tran, H. Wang, L. Torresani, J. Ray, Y. LeCun, and M. Paluri, “A Closer Look at Spatiotemporal Convolutions for Action Recognition,” 2018.

[33] W. Kay, J. Carreira, K. Simonyan, B. Zhang, C. Hillier, S. Vijayanarasimhan, F. Viola, T. Green, T. Back, P. Natsev, M. Suleyman, and A. Zisserman, “The Kinetics Human Action Video Dataset,” 2017.

[34] M. Raghu, C. Zhang, J. Kleinberg, and S. Bengio, “Transfusion: Understanding Transfer Learning for Medical Imaging,” 2019.

[35] B. Mustafa, A. Loh, J. Freyberg, P. MacWilliams, M. Wilson, S. M. McKinney, M. Sieniek, J. Winkens, Y. Liu, P. Bui, S. Prabhakara, U. Telang, A. Karthikesalingam, N. Houlsby, and V. Natarajan, “Supervised Transfer Learning at Scale for Medical Imaging,” 2021.

[36] Pérez-García, R. Sparks, and S. Ourselin, “TorchIO: a Python library for efficient loading, preprocessing, augmentation and patch-based sampling of medical images in deep learning,” Computer Methods and Programs in Biomedicine, p. 106236, 2021.

[37] D. P. Kingma and J. Ba, “Adam: A method for stochastic optimization,” arXiv preprint 1412.6980, 2014.

[38] J. Bergstra and Y. Bengio, “Random Search for Hyper-Parameter Optimization,” Journal of Machine Learning Research, vol. 13, no. 10, pp. 281–305, 2012.

[39] C. Szegedy, V. Vanhoucke, S. Ioffe, J. Shlens, and Z. Wojna, “Rethinking the inception architecture for computer vision,” in Proceedings of the IEEE conference on computer vision and pattern recognition, pp. 2818–2826, 2016.

[40] R. Müller, S. Kornblith, and G. Hinton, “When does label smoothing help?,” arXiv preprint 1906.02629, 2019.

[41] Huang, Y. Sun, Z. Liu, D. Sedra, and K. Q. Weinberger, “Deep networks with stochastic depth,” in European conference on computer vision, pp. 646–661, Springer, 2016.

[42] Y. Wu and K. He, “Group normalization,” in Proceedings of the European conference on computer vision (ECCV), pp. 3–19, 2018.

[43] S. Ioffe and C. Szegedy, “Batch normalization: Accelerating deep network training by reducing internal covariate shift,” in International conference on machine learning, pp. 448–456, PMLR, 2015.

[44] neptune.ai, “Neptune: experiment management and collaboration tool,” 2020.

[45] L. Biewald, “Experiment Tracking with Weights and Biases,” 2020. Software available from http://wandb.com.

[46] M. D. Herrmann, D. A. Clunie, F. A. S. W. Doyle, S. Pieper, V. Klepeis, L. P. Le, G. L. Mutter, D. S. Milstone, T. J. Schultz, R. Kikinis, G. K. Kotecha, D. H. Hwang, P. Andriole K, A. J. Iafrate, J. A. Brink, G. W. Boland, K. J. Dreyer, M. Michalski, J. A. Golden, D. N. Louis, and J. K. Lennerz, “Implementing the DICOM standard for digital pathology,” Journal of Pathology Informatics, vol. 9, 2018.

[47] T. Urban, E. Ziegler, R. Lewis, C. Hafey, C. Sadow, A. D. Van den Abbeele, and G. J. Harris, “LesionTracker: extensible open-source zero-footprint web viewer for cancer imaging research and clinical trials,” Cancer research, vol. 77, no. 21, pp. e119–e122, 2017.

[48] B. D. Gallas, H.-P. Chan, C. J. D’Orsi, L. E. Dodd, M. L. Giger, D. Gur, E. A. Krupinski, C. E. Metz, K. J. Myers, N. A. Obuchowski, et al., “Evaluating imaging and computer-aided detection and diagnosis devices at the FDA,” Academic radiology, vol. 19, no. 4, pp. 463–477, 2012.

[49] Y. Jiang, C. E. Metz, and R. M. Nishikawa, “A receiver operating characteristic partial area index for highly sensitive diagnostic tests.,” Radiology, vol. 201, no. 3, pp. 745–750, 1996.

[50] X. Robin, N. Turck, A. Hainard, N. Tiberti, F. Lisacek, J.-C. Sanchez, and M. Müller, “pROC: an open-source package for R and S+ to analyze and compare ROC curves,” BMC Bioinformatics, vol. 12, p. 77, 2011.

[51] J. Carpenter and J. Bithell, “Bootstrap confidence intervals: when, which, what? A practical guide for medical statisticians,” Statistics in medicine, vol. 19, no. 9, pp. 1141–1164, 2000.

[52] N. A. Obuchowski Jr and H. E. Rockette Jr, “Hypothesis testing of diagnostic accuracy for multiple readers and multiple tests an anova approach with dependent observations,” Communications in Statistics-simulation and Computation, vol. 24, no. 2, pp. 285–308, 1995.

[53] S. L. Hillis, N. A. Obuchowski, K. M. Schartz, and K. S. Berbaum, “A comparison of the dorfman–berbaum–metz and obuchowski–rockette methods for receiver operating characteristic (roc) data,” Statistics in medicine, vol. 24, no. 10, pp. 1579–1607, 2005.

[54] D. P. Chakraborty, Observer performance methods for diagnostic imaging: foundations, modeling, and applications with r-based examples. CRC Press, 2017.

[55] D. Chakraborty, P. Phillips, and X. Zhai, RJafroc: Artificial Intelligence Systems and Observer Performance, 2020. R package version 2.0.1.

[56] E. R. DeLong, D. M. DeLong, and D. L. Clarke-Pearson, “Comparing the areas under two or more correlated receiver operating characteristic curves: a nonparametric approach,” Biometrics, pp. 837–845, 1988.

[57] P. Capogrosso and A. J. Vickers, “A systematic review of the literature demonstrates some errors in the use of decision curve analysis but generally correct interpretation of findings,” Medical Decision Making, vol. 39, no. 5, pp. 493–498, 2019.

[58] K. F. Kerr, T. L. Marsh, and H. Janes, “The importance of uncertainty and opt-in v. opt-out: best practices for decision curve analysis,” 2019.

[59] J. L. Fleiss, “Measuring nominal scale agreement among many raters.,” Psychological bulletin, vol. 76, no. 5, p. 378, 1971.

[60] J. J. Randolph, “Free-Marginal Multirater Kappa (multirater K [free]): An Alternative to Fleiss’ Fixed-Marginal Multirater Kappa.,” Online submission, 2005.

[61] T. K. Koo and M. Y. Li, “A guideline of selecting and reporting intraclass correlation coefficients for reliability research,” Journal of chiropractic medicine, vol. 15, no. 2, pp. 155–163, 2016.

