## Supplementary Information for "Improving breast cancer diagnostics with artificial intelligence for MRI"

#### 1. Reader study results

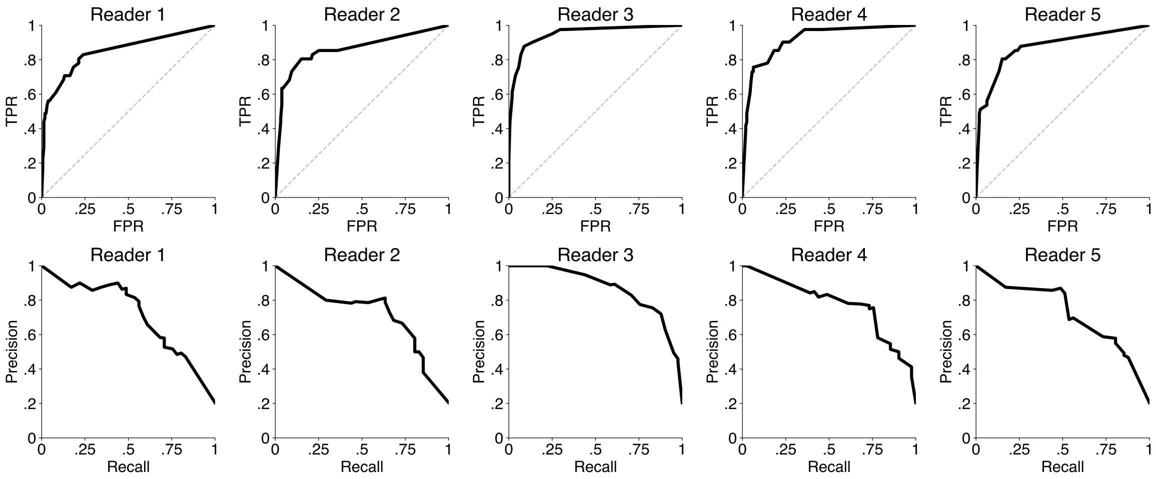


Figure S1: **All receiver operating characteristic (ROC) and precision-recall (PR) curves from the reader study**. The first two rows show ROC curves for all 5 readers, and the next two rows show PR curves. All ROC curves are non-parametric (empirical) and were generated from probabilities of malignancy provided by radiologists.

Table S1: **Reader study results**, reported with 95% confidence intervals estimated with bootstrap (N=2,000). We also report an average performance across all 5 readers. Studies classified by radiologists as BI-RADS category 4 or 5 were considered as positive and BI-RADS <4 as negative. Average reader performance was calculated as a simple mean of metrics for all readers. For AI predictions, a decision threshold was selected such that the AI system's sensitivity closely matches average reader sensitivity.

| Reader | AUROC | AUPRC | Sensitivity | Specificity | PPV | NPV |
|---|---|---|---|---|---|---|
| Reader 1 | 0.850 (0.779-0.917) | 0.712 (0.567-0.833) | 0.780 (0.659-0.895) | 0.786 (0.721-0.849) | 0.485 (0.381-0.593) | 0.933 (0.883-0.972) |
| Reader 2 | 0.860 (0.780-0.935) | 0.715 (0.556-0.853) | 0.854 (0.737-0.969) | 0.660 (0.582-0.745) | 0.393 (0.294-0.490) | 0.946 (0.904-0.991) |
| Reader 3 | 0.948 (0.908-0.978) | 0.868 (0.778-0.941) | 0.976 (0.913-1.000) | 0.704 (0.634-0.764) | 0.460 (0.366-0.556) | 0.991 (0.971-1.000) |
| Reader 4 | 0.916 (0.866-0.954) | 0.775 (0.640-0.867) | 0.976 (0.917-1.000) | 0.610 (0.536-0.679) | 0.392 (0.291-0.487) | 0.990 (0.965-1.000) |
| Reader 5 | 0.873 (0.820-0.932) | 0.721 (0.596-0.842) | 0.854 (0.750-0.949) | 0.761 (0.700-0.822) | 0.479 (0.370-0.582) | 0.953 (0.915-0.985) |
| Avg Reader | 0.890 | 0.758 | 0.888 | 0.704 | 0.442 | 0.962 |
| AI System | 0.924 (0.880-0.962) | 0.784 (0.656-0.887) | 0.897 (0.786-0.976) | 0.796 (0.728-0.856) | 0.517 (0.388-0.629) | 0.969 (0.937-0.993) |

#### 2. Hybrid predictions

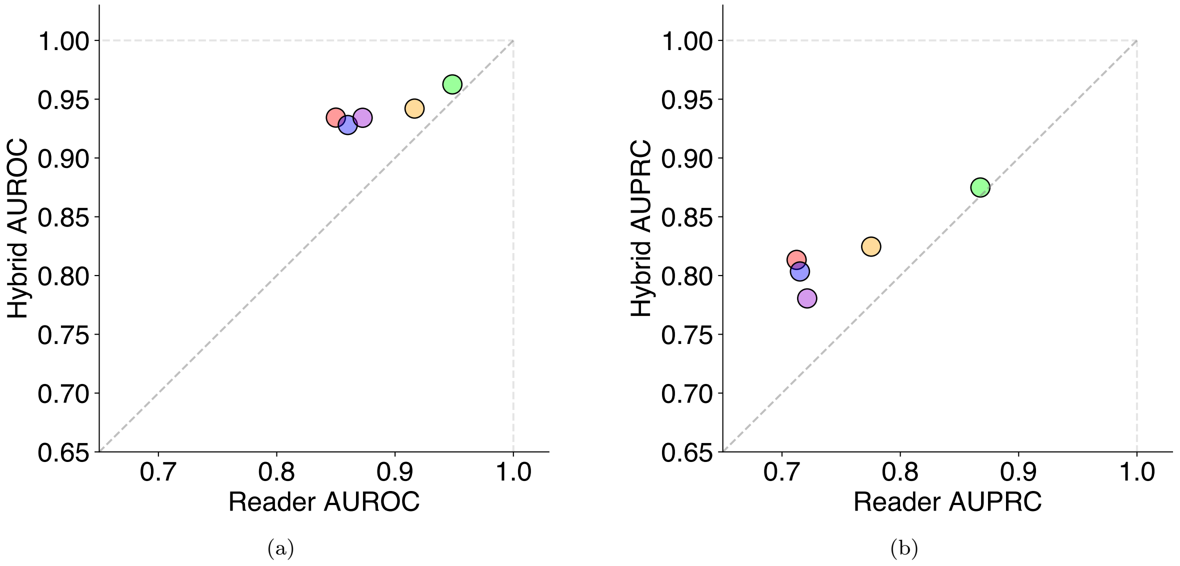


Figure S2: **Hybrid predictions are stronger than readers' predictions alone**. We demonstrate that an equally weighted average of radiologists and AI model predictions consistently yield a stronger performance in terms of both AUROC (**a**) and AUPRC (**b**). Performance increase is more marked in radiologists who performed slightly worse. However, even for the strongest reader's predictions the results are higher when averaged with AI model.

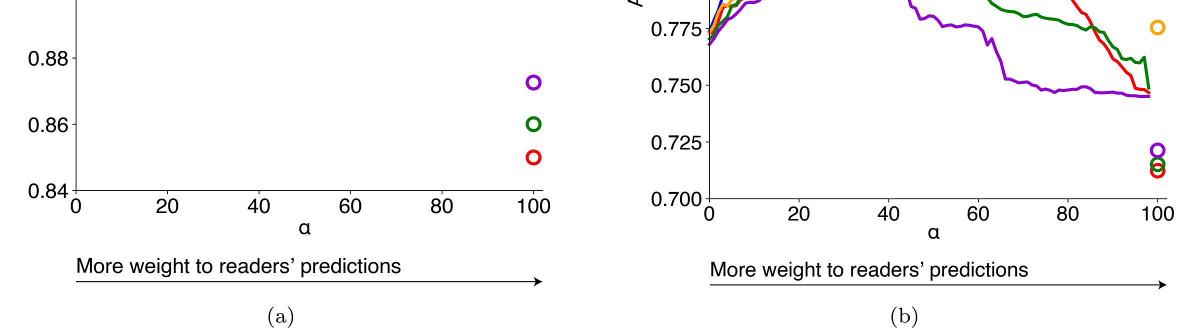


Figure S3: **Performance of a hybrid model, as a function of** $\alpha \in (0, 99]\%$**.** Plots show how **a**, AUROC and **b**, AUPRC change when the $\alpha$ multiplier changes. At $\alpha = 0\%$, the hybrid performance is equal to the model only performance. At $\alpha = 100\%$, the hybrid performance is equal to the reader only performance (here plotted as a point on the far right of the figures). Results demonstrate that utilizing AI predictions even at low weights (high $\alpha$) significantly improves the performance. Each line represents performance for a different reader. Diamond-shaped points represent maximum performance for each metric and reader.

#### 3. Subgroup performance

Table S2: **Subgroup performance.** Reported values are $N$ (95% confidence intervals). Confidence intervals were calculated with a bootstrap (2,000 replicates). $PPV$, positive predictive value; $NPV$, negative predictive value. As there were no malignant examples in BI-RADS 1 and 2 categories in our test set, AUROC would not be defined for those groups. BI-RADS 1 and 2 were combined with BI-RADS 3 patients to generate the results. For AI predictions, a decision threshold was selected such that the AI system's sensitivity closely matches average reader sensitivity.

| Group | $N$ | AUROC | AUPRC | Sensitivity | Specificity | PPV | NPV |
|---|---|---|---|---|---|---|---|
| BIRADS | | | | | | | |
| BIRADS 1/2/3 | 2,307 | 0.84 (0.68-0.97) | 0.09 (0.01-0.27) | 0.75 (0.46-1.00) | 0.83 (0.82-0.84) | 0.01 (0.00-0.02) | 1.00 (1.00-1.00) |
| BIRADS 4 | 956 | 0.87 (0.85-0.89) | 0.72 (0.67-0.76) | 0.88 (0.85-0.91) | 0.61 (0.58-0.63) | 0.42 (0.39-0.45) | 0.94 (0.93-0.96) |
| BIRADS 5 | 40 | 0.87 (0.78-0.95) | 0.89 (0.78-0.97) | 0.91 (0.82-0.98) | 0.47 (0.31-0.64) | 0.68 (0.56-0.79) | 0.81 (0.64-0.95) |
| BIRADS 6 | 385 | 0.90 (0.87-0.92) | 0.88 (0.85-0.92) | 0.90 (0.86-0.93) | 0.67 (0.62-0.72) | 0.70 (0.65-0.74) | 0.88 (0.85-0.92) |
| BIRADS 0 | 102 | 0.94 (0.88-0.98) | 0.61 (0.35-0.84) | 0.94 (0.82-1.00) | 0.75 (0.68-0.81) | 0.27 (0.16-0.38) | 0.99 (0.98-1.00) |
| unknown | 146 | 0.92 (0.87-0.96) | 0.75 (0.61-0.85) | 0.89 (0.81-0.98) | 0.79 (0.74-0.84) | 0.45 (0.35-0.55) | 0.97 (0.95-0.99) |
| Age | | | | | | | |
| Age $<40$ | 399 | 0.91 (0.87-0.94) | 0.65 (0.53-0.76) | 0.89 (0.81-0.95) | 0.74 (0.70-0.77) | 0.27 (0.22-0.33) | 0.98 (0.97-0.99) |
| Age $<50$ | 1,294 | 0.91 (0.89-0.93) | 0.67 (0.61-0.73) | 0.89 (0.85-0.92) | 0.73 (0.71-0.75) | 0.30 (0.27-0.33) | 0.98 (0.97-0.99) |
| Age $\geq 50$ | 2,642 | 0.93 (0.92-0.94) | 0.74 (0.71-0.78) | 0.89 (0.86-0.91) | 0.79 (0.78-0.80) | 0.37 (0.35-0.40) | 0.98 (0.98-0.99) |
| Histology | | | | | | | |
| DCIS | 570 | 0.91 (0.89-0.92) | 0.93 (0.92-0.95) | 0.89 (0.86-0.91) | 0.68 (0.64-0.72) | 0.76 (0.72-0.79) | 0.84 (0.80-0.87) |
| IDC | 523 | 0.93 (0.92-0.95) | 0.95 (0.94-0.96) | 0.92 (0.90-0.94) | 0.64 (0.60-0.68) | 0.74 (0.71-0.78) | 0.88 (0.84-0.91) |
| Meta | 138 | 0.96 (0.93-0.98) | 0.97 (0.94-0.99) | 0.96 (0.93-0.99) | 0.58 (0.49-0.66) | 0.72 (0.66-0.79) | 0.93 (0.86-0.98) |
| Adenoca | 106 | 0.95 (0.92-0.98) | 0.96 (0.93-0.98) | 0.98 (0.95-1.00) | 0.55 (0.45-0.65) | 0.72 (0.65-0.79) | 0.96 (0.91-1.00) |
| ILC | 87 | 0.90 (0.85-0.94) | 0.94 (0.90-0.96) | 0.86 (0.79-0.92) | 0.63 (0.52-0.74) | 0.75 (0.67-0.83) | 0.77 (0.67-0.87) |
| IMC | 33 | 0.94 (0.88-0.99) | 0.95 (0.89-0.99) | 0.94 (0.85-1.00) | 0.75 (0.59-0.90) | 0.80 (0.67-0.92) | 0.92 (0.81-1.00) |
| Other/unknown | 20 | 0.84 (0.70-0.96) | 0.91 (0.79-0.98) | 0.81 (0.63-0.95) | 0.53 (0.29-0.75) | 0.65 (0.46-0.83) | 0.71 (0.45-0.93) |
| Molecular | | | | | | | |
| Luminal A | 326 | 0.93 (0.90-0.94) | 0.95 (0.93-0.96) | 0.91 (0.88-0.94) | 0.68 (0.63-0.73) | 0.77 (0.73-0.81) | 0.86 (0.82-0.90) |
| Luminal B | 78 | 0.96 (0.92-0.99) | 0.97 (0.94-0.99) | 0.96 (0.91-1.00) | 0.67 (0.56-0.78) | 0.76 (0.67-0.84) | 0.94 (0.87-1.00) |
| Triple negative | 63 | 0.93 (0.87-0.97) | 0.95 (0.91-0.98) | 0.91 (0.82-0.97) | 0.71 (0.59-0.83) | 0.76 (0.66-0.86) | 0.88 (0.78-0.96) |
| HER2-enriched | 21 | 0.97 (0.91-1.00) | 0.98 (0.93-1.00) | 0.95 (0.85-1.00) | 0.67 (0.45-0.85) | 0.74 (0.57-0.89) | 0.93 (0.79-1.00) |
| BPE | | | | | | | |
| Minimal | 884 | 0.94 (0.92-0.96) | 0.78 (0.71-0.84) | 0.89 (0.84-0.94) | 0.85 (0.83-0.86) | 0.35 (0.30-0.39) | 0.99 (0.98-0.99) |
| Mild | 1,614 | 0.93 (0.91-0.94) | 0.72 (0.68-0.77) | 0.89 (0.86-0.92) | 0.79 (0.77-0.80) | 0.37 (0.34-0.40) | 0.98 (0.97-0.99) |
| Moderate | 884 | 0.91 (0.88-0.93) | 0.71 (0.65-0.77) | 0.88 (0.84-0.92) | 0.68 (0.66-0.71) | 0.32 (0.28-0.35) | 0.97 (0.96-0.98) |
| Marked | 184 | 0.87 (0.82-0.92) | 0.66 (0.54-0.77) | 0.92 (0.85-0.98) | 0.60 (0.54-0.65) | 0.33 (0.27-0.40) | 0.97 (0.95-0.99) |
| Unknown | 370 | 0.92 (0.88-0.95) | 0.67 (0.56-0.77) | 0.86 (0.78-0.94) | 0.79 (0.76-0.82) | 0.33 (0.27-0.39) | 0.98 (0.97-0.99) |
| Race | | | | | | | |
| White | 2,738 | 0.93 (0.91-0.94) | 0.72 (0.68-0.75) | 0.88 (0.85-0.90) | 0.78 (0.77-0.80) | 0.32 (0.30-0.34) | 0.98 (0.98-0.99) |
| Black | 244 | 0.91 (0.87-0.94) | 0.82 (0.75-0.89) | 0.88 (0.82-0.94) | 0.73 (0.69-0.78) | 0.49 (0.43-0.57) | 0.95 (0.93-0.98) |
| Asian | 163 | 0.94 (0.89-0.97) | 0.82 (0.71-0.93) | 0.92 (0.85-0.98) | 0.71 (0.65-0.76) | 0.44 (0.36-0.52) | 0.97 (0.95-0.99) |
| Other Race | 237 | 0.90 (0.86-0.93) | 0.71 (0.60-0.81) | 0.90 (0.83-0.95) | 0.67 (0.62-0.72) | 0.40 (0.34-0.47) | 0.96 (0.94-0.98) |
| Magnet | | | | | | | |
| 1.5T | 2,102 | 0.93 (0.91-0.94) | 0.67 (0.62-0.72) | 0.84 (0.80-0.87) | 0.86 (0.85-0.87) | 0.36 (0.32-0.39) | 0.98 (0.98-0.99) |
| 3T | 1,834 | 0.92 (0.91-0.93) | 0.75 (0.72-0.79) | 0.92 (0.90-0.94) | 0.66 (0.65-0.68) | 0.34 (0.32-0.36) | 0.98 (0.97-0.98) |

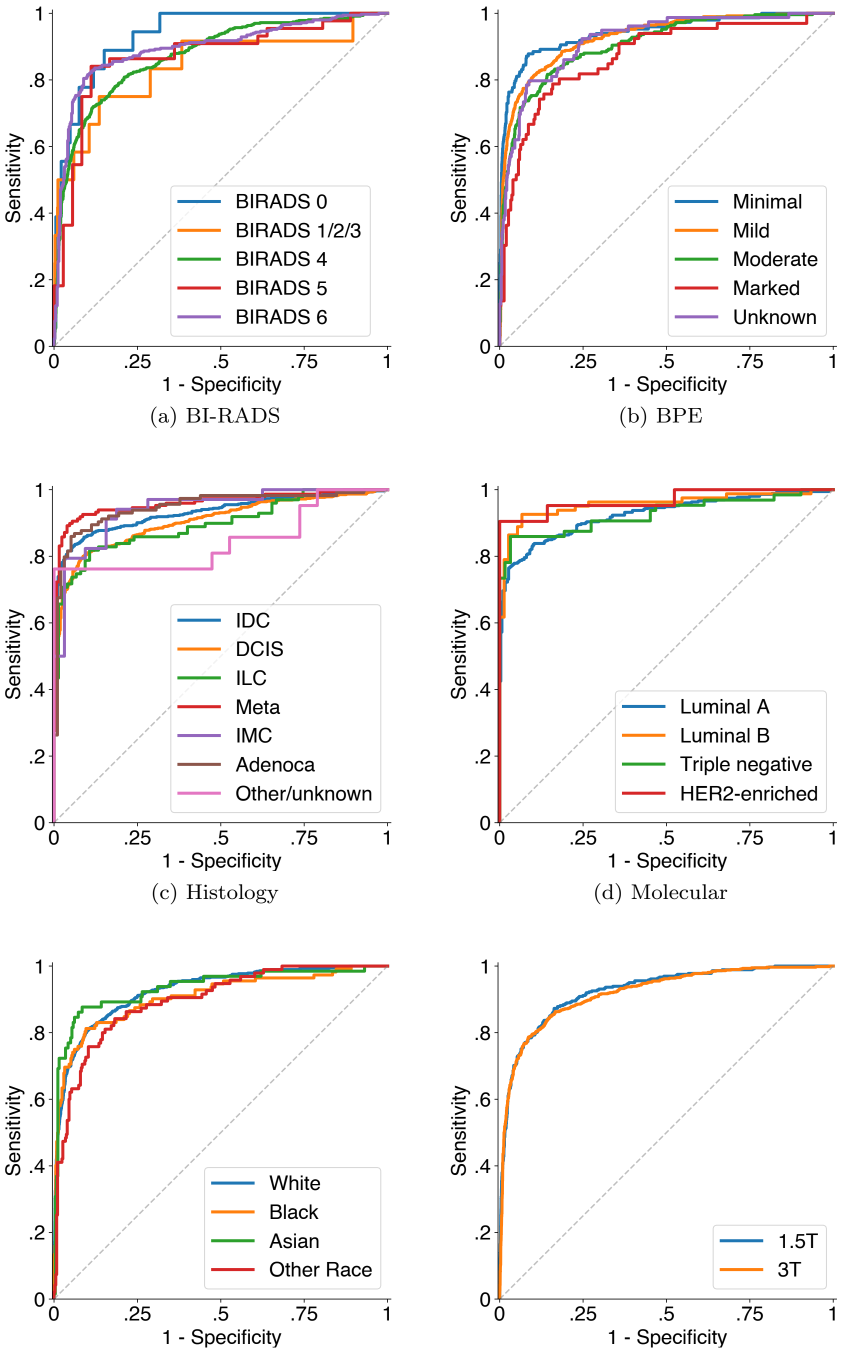


(a) BI-RADS

(b) BPE

(c) Histology

(d) Molecular

(e) Race

(f) Magnet strength

Figure S4: **Empirical ROC curves for subgroups** per BI-RADS risk category, background parenchymal enhancement category, histological subtype, molecular subtype, race, and magnet strength. As there were no malignant examples in BI-RADS 1 and 2 categories in our test set, AUC ROC would not be defined for those groups. BI-RADS 1 and 2 were combined with BI-RADS 3 patients to generate AUC ROC and curves.

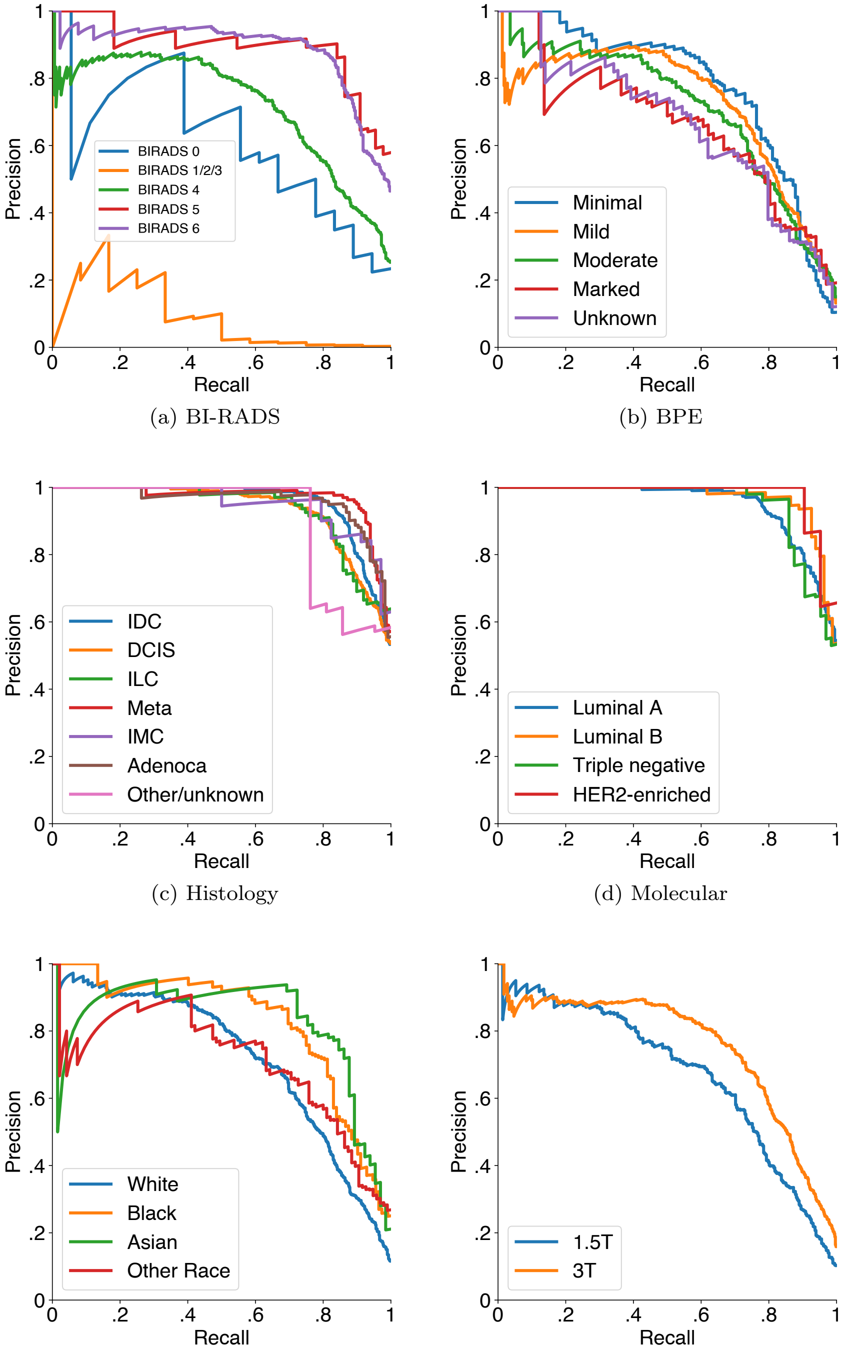


(a) BI-RADS (b) BPE

(c) Histology (d) Molecular

(e) Race (f) Magnet strength

Figure S5: **Empirical precision-recall curves for subgroups** per BI-RADS risk category, background parenchymal enhancement category, histological subtype, molecular subtype, race and magnet strength. As there were no malignant examples in BI-RADS 1 and 2 categories in our test set, AUC PR would not be defined for those groups. BI-RADS 1 and 2 were combined with BI-RADS 3 patients to generate AUC PR and curves.

### 4. BI-RADS downgrading

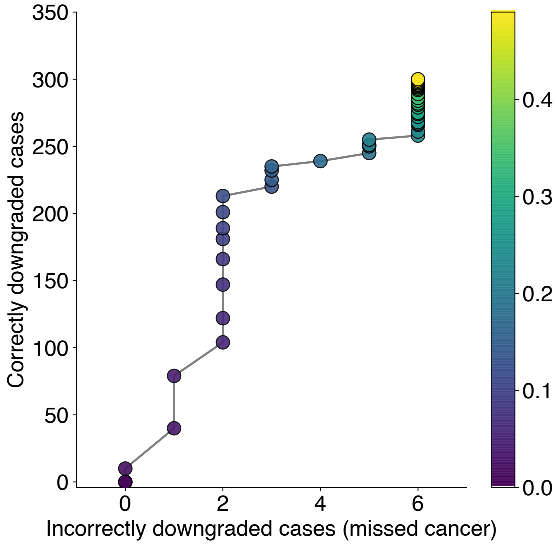


(a) BI-RADS 3

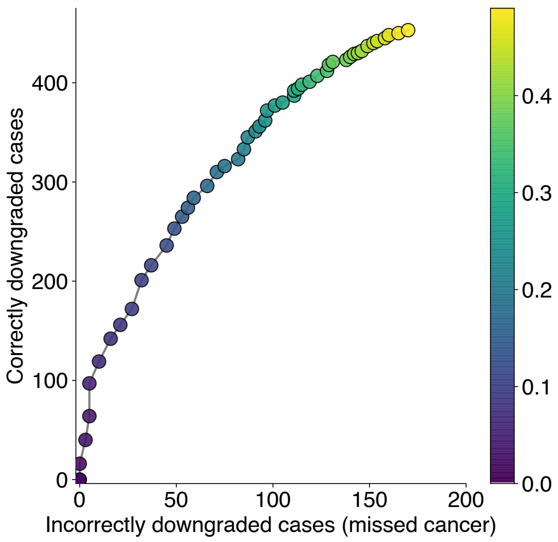


(b) BI-RADS 4

Figure S6: **Trade-off in missed cancers versus correctly avoided interventions when using only AI system to decide on management.** **a**, shows the trade-off when using only AI system's predictions to decide whether the patient should return for a 6-month follow-up or not in BI-RADS 3 cases. "Correctly downgraded" patients would return to a regular screening, while "missed cancers" prevent the opportunity to detect cancer if it would be imaged again in 6 months. **b**, shows the trade-off in BI-RADS 4 cases. "Correctly downgraded" cases from BI-RADS 4 to BI-RADS 3 represent patients who would avoid an unnecessary biopsy, while "missed cancers" are situations where patients do have breast cancer but would not be biopsied because of the AI system's predictions. Both **a,b** do not take into consideration patient's and physician's preferences and do not weigh the trade-off items (e.g. one missed cancer case is more important than one avoided biopsy). They also ignore the potential effect of physician ultimately making a decision based on their own knowledge supported by the AI system. **a,b** show the trade-off at different operating points. Operating points are color-coded by increasing binarization thresholds (warmer colors are higher thresholds).

#### 5. Processing TCGA-BRCA data set

In its original form, TCGA-BRCA data set is not suitable for AI evaluation or training purposes. Specifically, it:

- contains studies where series for left and right breasts are separated,
- contains studies where one or more series are multi-volume,
- contains studies where only one breast is imaged,
- does not provide information on which series are pre- and post-contrast,
- does not provide breast-level labels.

To solve this problem, we established a pipeline for processing the TCGA-BRCA data set for AI purposes. This means that the script we share in our manuscript repository [1] is able to take a downloaded data set in its current form and return NIfTI files for pre- and two post-contrast series. For series where two breasts are saved separately, the script merges them into a single volume. For series where both breasts are imaged, but multiple acquisitions are saved in a single volume (multi-volume), the script splits the multi-volume into separate series. Additionally, the script excludes studies that are unilateral. Along with the script, we provide a YAML file which defines a list of all TCGA-BRCA studies for inclusion/exclusion, type of laterality and, potential problems (multi-volumes etc.) as well as series numbers corresponding to pre- and post-contrast T1 fat-sat series.

Labels for the TCGA-BRCA data set have been generated using one of the supporting files (`clinical_patient_brca.txt`), specifically `anatomic_neoplasm_subdivision` column.

[1] `https://github.com/nyukat/MRI_AI`

#### 6. Manufacturers and devices

Device names were acquired by extracting information from DICOM tags `Manufacturer` and `ManufacturerModelName`. For Duke University data set, this information was collected from a spreadsheet provided by data set authors and available at The Cancer Imaging Archive (file "Clinical and Other Features").

In total, there were:

- 21,602 studies acquired on Siemens machines,
- 840 studies acquired on GE machines,
- 493 studies acquired on Philips machines,
- 24 studies acquired on Hitachi machines,
- 28 studies with missing information about the scanner.

Table S3: **MRI manufacturer and model breakdown for all data sets.** MRI scanners are sorted by number of total cases in the data set, descending. If a cell is empty, that means that the specific data set does not contain any cases acquired on the machine. †In 28 unknown cases, manufacturer information was not provided and DICOM tags were populated with the hospital PACS name.

| | | ***Number of cases*** | | | | |
|---|---|---|---|---|---|---|
| **Manufacturer/Model** | **Magnet** | **NYU Langone** | **Jagiellonian University** | **Duke University** | **TCGA-BRCA** | **Total** |
| Siemens Symphony | 1.5T | 9,638 | | | 2 | 9,640 |
| Siemens Trio Tim | 3T | 8,142 | | 58 | | 8,200 |
| Siemens Skyra | 3T | 1,940 | 1 | 57 | | 1,998 |
| Siemens Espree | 1.5T | 668 | | | 1 | 669 |
| Philips Achieva | 1.5T | 477 | | | 16 | 493 |
| Siemens MAGNETOM Sola | 1.5T | | 395 | | | 395 |
| Siemens Avanto | 1.5T | 132 | 1 | 179 | 3 | 315 |
| GE SIGNA HDx | 1.5T | | | 272 | 8 | 280 |
| GE SIGNA HDxt | 1.5T | | | 248 | 6 | 254 |
| Siemens Verio | 3T | 175 | | | | 175 |
| GE SIGNA HDe | 1.5T | 112 | | | | 112 |
| Siemens Aera | 1.5T | 72 | | | | 72 |
| Siemens Verio Dot | 3T | 65 | | | | 65 |
| Siemens MAGNETOM Vida | 3T | 64 | | | | 64 |
| Hitachi ECHELON | 1.5T | 24 | | | | 24 |
| GE Optima MR450w | 1.5T | | | 98 | | 98 |
| GE SIGNA EXCITE | 1.5T | | | 10 | 85 | 95 |
| Siemens Sonata | 3T | | | | 9 | 9 |
| GE DISCOVERY MR750 | 3T | | | | 1 | 1 |
| Unknown† | - | 28 | | | | 28 |
| | | **21,537** | **397** | **922** | **131** | **22,987** |

#### 7. Breast-level labels

Table S4: **Breast-level breakdown of labels in the NYU Langone data set.** Malignant and benign labels are not mutually exclusive. A patient might have both a malignant and a benign change in the same breast.

| | Training set | Validation set | Test set | Total |
|---|---|---|---|---|
| **Breast-level labels** | | | | |
| Left benign | 2,117 | 518 | 715 | 3,350 |
| Right benign | 2,111 | 477 | 705 | 3,293 |
| Left malignant | 1,278 | 326 | 478 | 2,082 |
| Right malignant | 1,211 | 293 | 427 | 1,931 |
| Left negative | 11,539 | 2,747 | 2,992 | 17,278 |
| Right negative | 11,617 | 2,798 | 3,060 | 17,475 |

#### 8. Distribution of predicted probabilities of malignancy

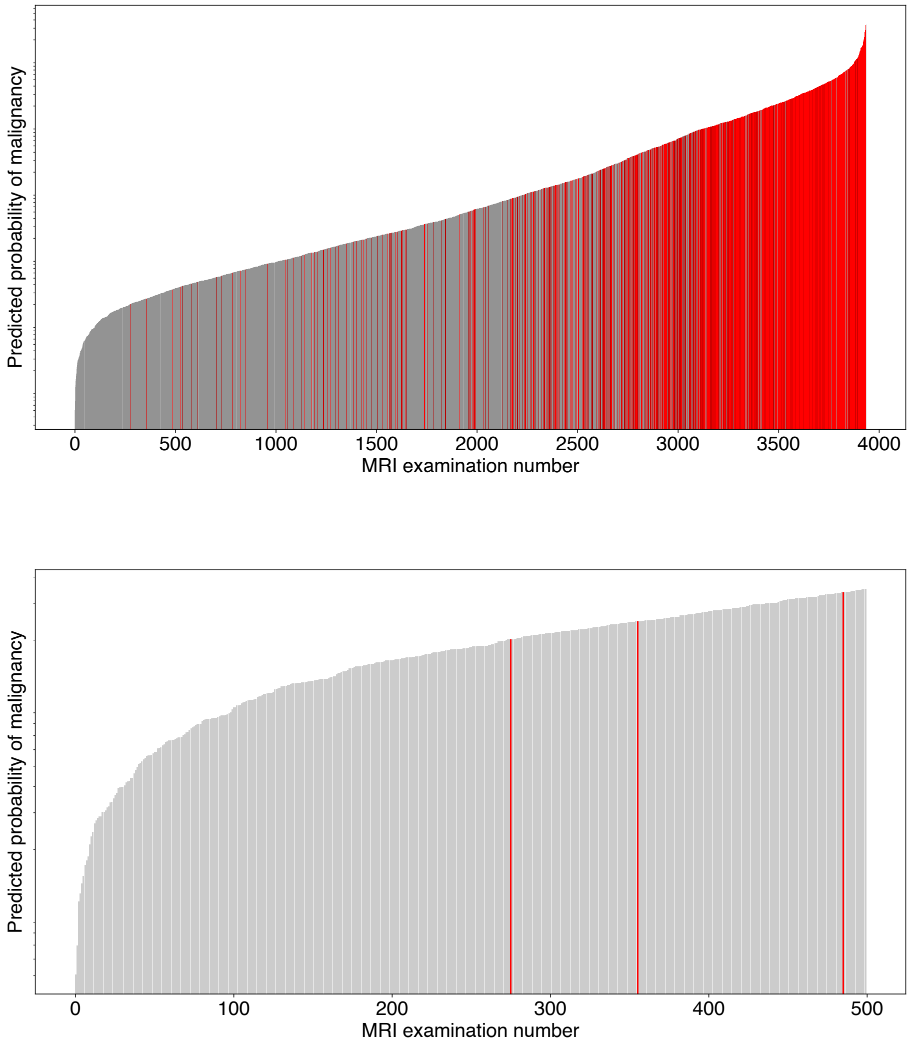


Figure S7: **Distribution of predicted probabilities of malignancy (POM) on the NYU Langone test set.** Each bar represents a POM for a single study (maximum between left and right breast POMs) and all bars are ordered by POM in an increasing manner. Red bars represent malignant cases, while black bars are non-malignant. **Top figure** shows all NYU Langone test set cases, meanwhile **bottom figure** zooms in on the first 500 cases.

### 9. Error analysis

Below are several studies, selected from the reader study subset, that show situations where our AI system is compared with radiologists predictions. We present probabilities of malignancy (POMs) for all readers and the AI system with a short case description.

#### 9.1. Correctly identified cancers

**Case 1.** In the following study, all five radiologists gave it a very high probability of malignancy in the right breast (one BI-RADS 4C, four BI-RADS 5). The AI system also correctly identified the malignancy and gave the study a 97% probability of cancer in the right breast. Interestingly, one radiologist found a suspicious lesion in the left breast. Based on the patients' history, that lesion was also identified by the radiologist originally interpreting the study. Upon biopsy, the lesion was found to be benign.

| | Left breast POM | Right breast POM |
|---|---|---|
| Reader 1 | 0 | 100 |
| Reader 2 | 0 | 99 |
| Reader 3 | 0 | 98 |
| Reader 4 | 10 | 99 |
| Reader 5 | 0 | 90 |
| **AI system** | **2** | **97** |

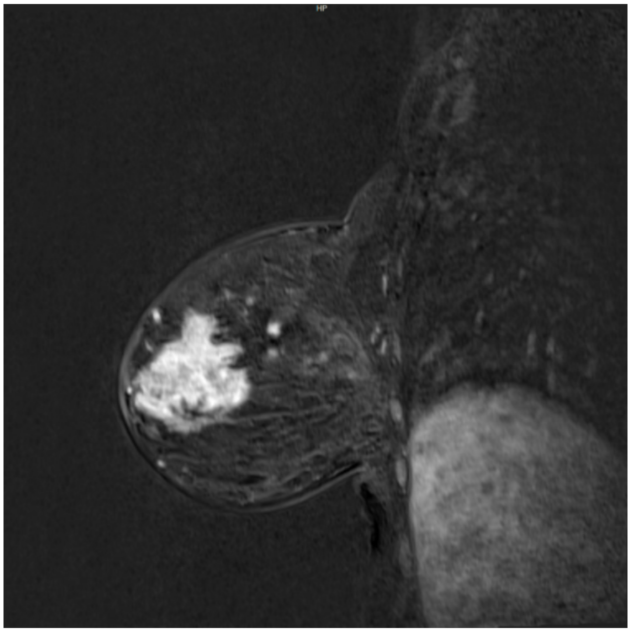

Figure S8: Sagittal view of the adenocarcinoma in the right breast. There are multiple irregular heterogeneously enhancing masses suspicious for satellite lesions.

**Case 2.** Here, only three out of five readers found any lesions in the study. Out of the three who did, only one gave it a high probability of malignancy (reader 5, 30%). The suspicious lesion was later confirmed to be malignant. Our AI model correctly predicted the malignancy, giving a 39% probability in the left breast, and 0% POM in the right breast.

| | Left breast POM | Right breast POM |
|---|---|---|
| Reader 1 | 2 | 0 |
| Reader 2 | 0 | 0 |
| Reader 3 | 5 | 0 |
| Reader 4 | 0 | 0 |
| Reader 5 | 30 | 2 |
| **AI system** | **39** | **0** |

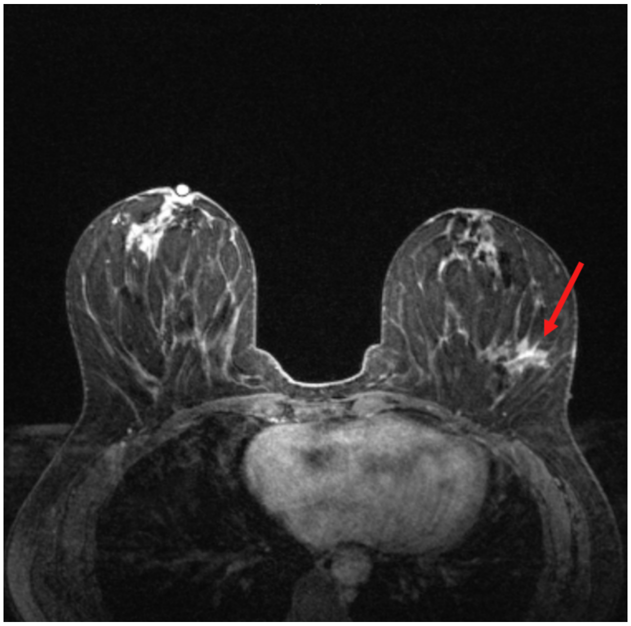

Figure S9: From the radiology report: "A 2 cm biopsy tract [red arrow] is present in the left outer breast at 3:00 posterior depth, associated with mild inflammatory changes and a biopsy clip in its medial aspect, concordant with the site of biopsy-proven malignancy."

**Case 3.** This study was performed in the diagnostic process of evaluating bloody left nipple discharge which demonstrated atypical cells. While there were no suspicious findings in the left breast, all radiologists agreed that the enhancement in the right breast is highly suspicious. This prediction was matched by AI output. Lesion was found to be malignant.

| | Left breast POM | Right breast POM |
|---|---|---|
| Reader 1 | 0 | 85 |
| Reader 2 | 0 | 50 |
| Reader 3 | 0 | 40 |
| Reader 4 | 0 | 85 |
| Reader 5 | 0 | 95 |
| **AI system** | **0** | **26** |

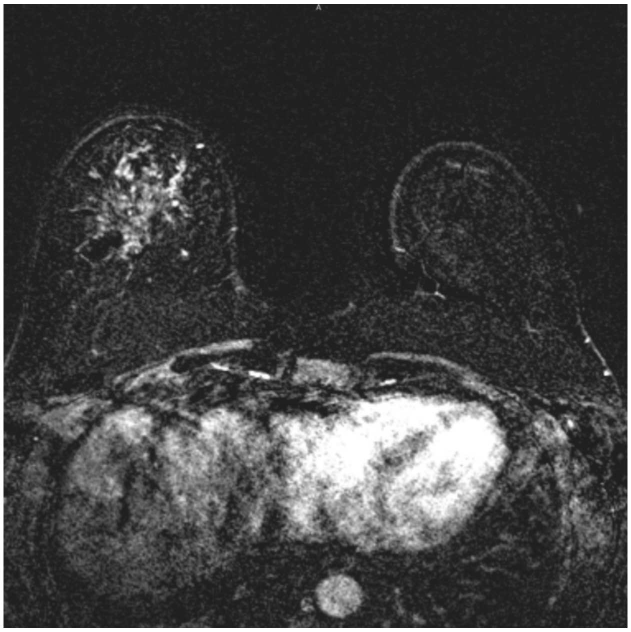

Figure S10: A slide from T1-weighted subtraction series with visible suspicious lesion in the right breast. From the radiology report: "Extensive nonmass enhancement in the inferior right breast with questionable mild architectural distortion".

#### 9.2. Correctly identified negative studies

**Case 4.** and **Case 5.** Below are two sample studies where all radiologists agreed that there are no suspicious lesions in the study, and our AI system gave very low probabilities of malignancy as well. Predictions in the table below are appropriate for both Case 4 and Case 5.

| | Left breast POM | Right breast POM |
|---|---|---|
| Reader 1 | 0 | 0 |
| Reader 2 | 0 | 0 |
| Reader 3 | 0 | 0 |
| Reader 4 | 0 | 0 |
| Reader 5 | 0 | 0 |
| **AI system** | **1** | **1** |

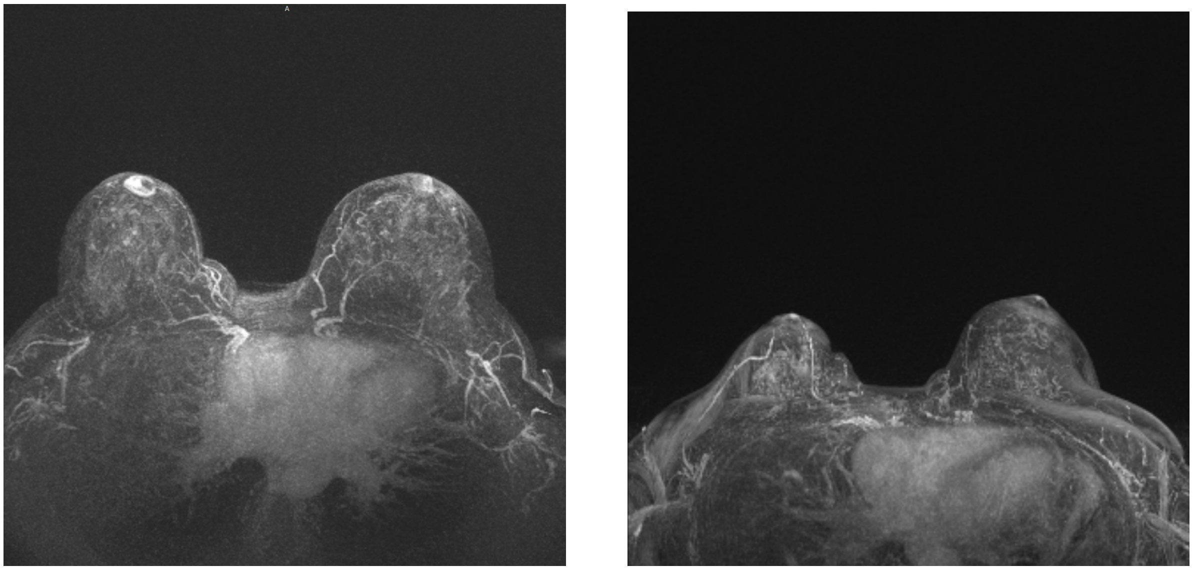

(a) (b)

Figure S11: Maximum intensity projection images for Case 4 (a) and Case 5 (b).

#### 9.3. Opportunity to avoid biopsies

**Case 6.** shows a study where all radiologists would biopsy the lesion in the left breast. One reader gave this exam BI-RADS 4A, three 4B and one 4C classification. Looking into patient history, the suspicious lesion in the left breast was indeed biopsied and yielded a benign result. Our AI system correctly outputted a low POM. This raises questions whether radiologists would be more likely to revisit their first diagnosis when provided with AI output.

| | Left breast POM | Right breast POM |
|---|---|---|
| Reader 1 | 15 | 0 |
| Reader 2 | 10 | 0 |
| Reader 3 | 50 | 0 |
| Reader 4 | 2 | 0 |
| Reader 5 | 10 | 0 |
| **AI system** | **1** | **0** |

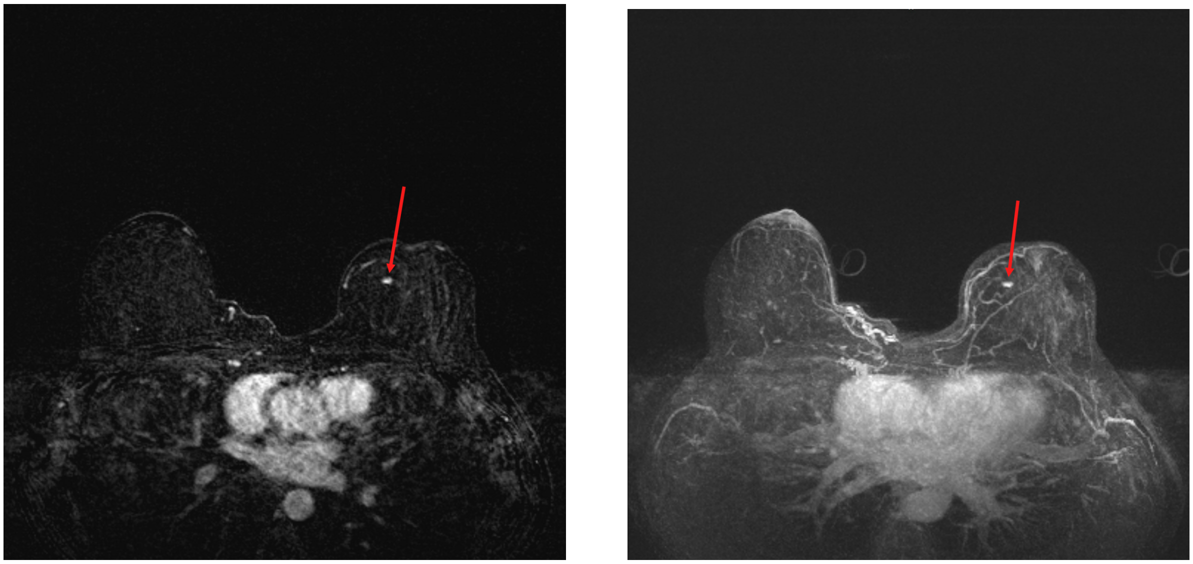

(a) (b)

Figure S12: Axial view of a subtraction image (a) and maximum intensity projection (b) of the Case 6 with visible lesion [red arrow] that was interpreted as suspicious by radiologists, but turned out to be benign after a core biopsy. Diagnosis from the pathology report said: "benign breast tissue with dense stroma, focal sclerosing adenosis, benign adipose tissue".

#### 9.4. Missed cancers

In this section we will investigate a few situations where patients were diagnosed with breast cancer, but our AI system output suggested low or very low probability of malignancy. We identified two studies where our system dramatically underestimated the POM (Case 7. and Case 8.). We also present two more studies where POM was higher, but still lower than preferable.

**Case 7.** Here, all radiologists agreed that right breast has a high POM with BI-RADS 4C/5. This was a situation where our model failed completely, yielding only 1% POM for the right breast. This study was performed to evaluate the extent of disease.

| | Left breast POM | Right breast POM |
|---|---|---|
| Reader 1 | 0 | 75 |
| Reader 2 | 0 | 99 |
| Reader 3 | 0 | 80 |
| Reader 4 | 0 | 95 |
| Reader 5 | 0 | 70 |
| **AI system** | **2** | **1** |

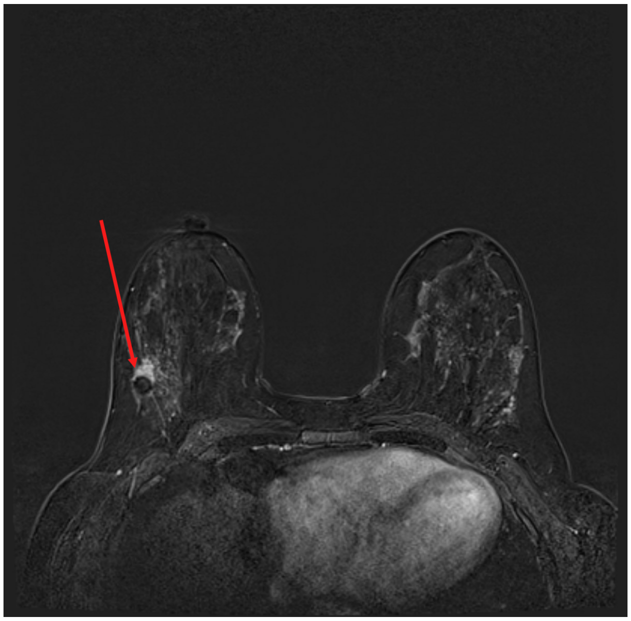

Figure S13: From radiology report: "2.5 x 1.6 x 2.2 cm enhancing mass containing susceptibility artifact from biopsy marker clip in the right breast 9:00 axis, 8 cm from the nipple, biopsy proven malignancy".

**Case 8.** Similarly to Case 7., all radiologists agreed that the left breast has a relatively high POM. Study was performed for extent of disease.

| | Left breast POM | Right breast POM |
|---|---|---|
| Reader 1 | 20 | 0 |
| Reader 2 | 50 | 0 |
| Reader 3 | 20 | 0 |
| Reader 4 | 40 | 0 |
| Reader 5 | 75 | 0 |
| **AI system** | **2** | **1** |

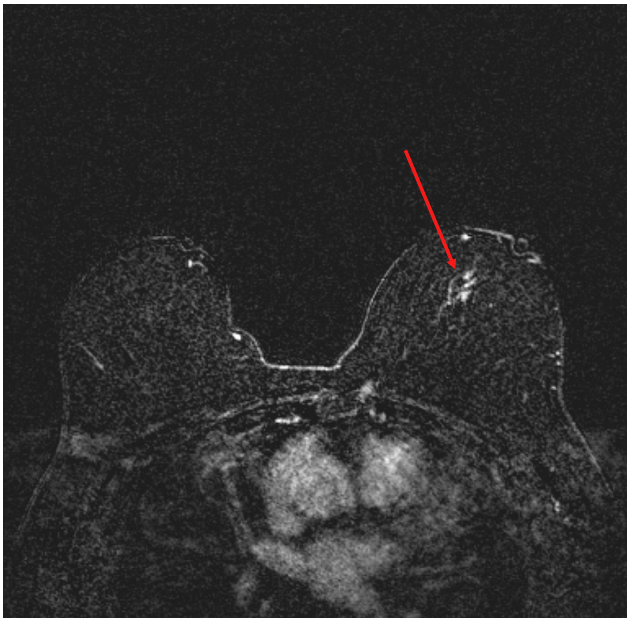

Figure S14: From radiology report: "Susceptibility artifact from a metallic clip is seen in the left breast mid depth with surrounding non mass enhancement collectively measuring 1.5 x 2.6 cm consistent with biopsy proven malignancy". Suspicious area marked with the red arrow.

**Case 9.** In this case, there were multiple suspicious findings in the right breast. Both radiologists and our AI system identified higher-than-average POM. However, the AI's POM was lower than expected from a highly accurate system. On the other hand, this POM was on par with some radiologists' predictions. Reader 1 would not even perform a biopsy, and Reader 4 gave a 10% POM for the right breast, the same that the AI system did.

| | Left breast POM | Right breast POM |
|---|---|---|
| Reader 1 | 0 | 1 |
| Reader 2 | 0 | 30 |
| Reader 3 | 0 | 50 |
| Reader 4 | 0 | 10 |
| Reader 5 | 0 | 85 |
| **AI system** | **2** | **10** |

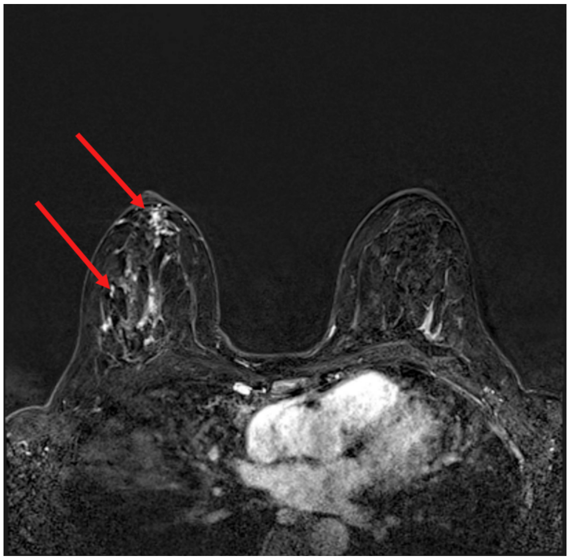

(a)

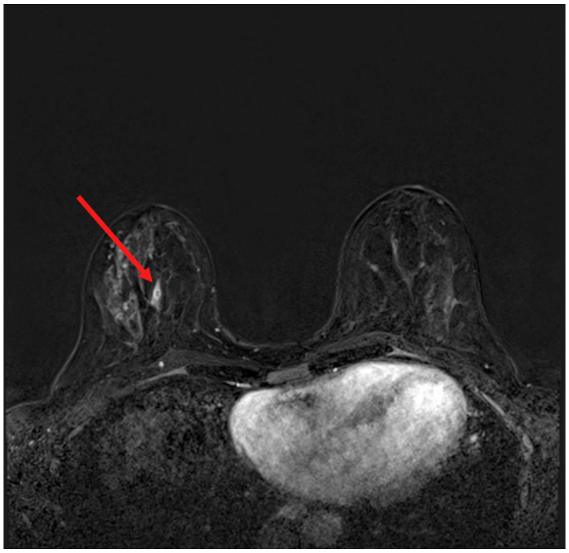

(b)

Figure S15: Axial subtraction images of multiple suspicious findings identified by radiologists, marked with red arrows. Two specimens were obtained in the core biopsy following the MRI, and they both yielded ductal carcinoma in situ (high nuclear grade, solid and cribriform types, with necrosis and focal microcalcifications).

#### 9.5. Overestimated POM on negative/benign cases

Here we investigate a few situations where the AI system outputted a high probability of malignancy, even though the case turned out to be benign or negative.

**Case 10.** Here, four out of five radiologists interpreted the study as negative. One radiologist (Reader 1) would biopsy the right breast. Our system, surprisingly, gave a relatively high POM for the right breast (68%).

| | Left breast POM | Right breast POM |
|---|---|---|
| Reader 1 | 0 | 5 |
| Reader 2 | 0 | 0 |
| Reader 3 | 0 | 0 |
| Reader 4 | 0 | 0 |
| Reader 5 | 0 | 0 |
| **AI system** | **6** | **68** |

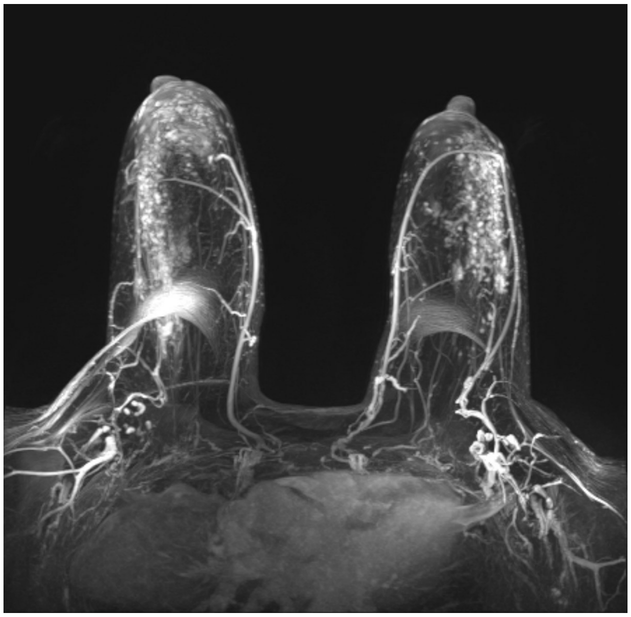

Figure S16: Axial maximum intensity projection from Case 10.

**Case 11** In this study, four out of five radiologists would biopsy the finding in right breast, and POM given to this study varied significantly. Ultimately, the finding was biopsied and was found benign. While our system's POM was very similar to radiologists, we would expect a highly accurate model to give lower POM to benign cases.

| | Left breast POM | Right breast POM |
|---|---|---|
| Reader 1 | 0 | 0 |
| Reader 2 | 0 | 75 |
| Reader 3 | 0 | 10 |
| Reader 4 | 0 | 30 |
| Reader 5 | 0 | 75 |
| **AI system** | **4** | **55** |

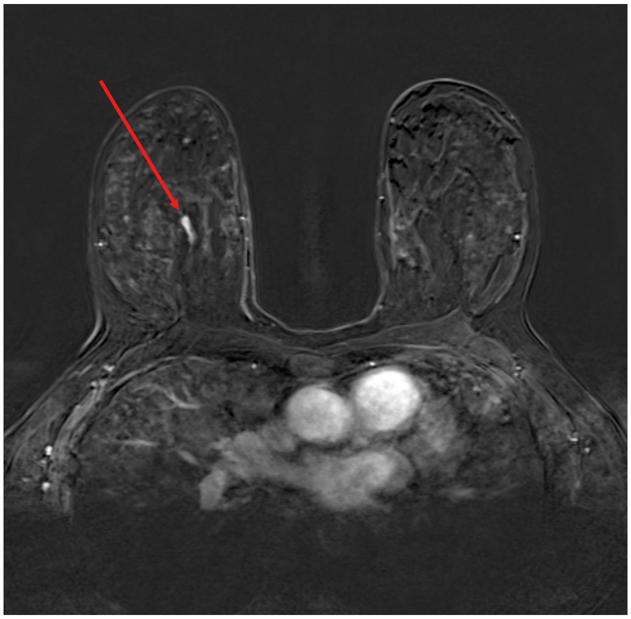

Figure S17: Axial subtraction image showing suspicious lesion that was later biopsied. Pathology report showed that the finding was benign, yielding fibrocystic changes, including columnar changes and stromal fibrosis.
